# Association between proximity to a lead releasing facility and cognition in diverse cohorts

**DOI:** 10.1101/2025.07.27.25332267

**Authors:** Scarlet Cockell, Kelly M. Bakulski, Ai-Lin Tsai, Yike Liu, Amanda J. Goodrich, Chinomnso Okorie, Stacey Alexeeff, Rachel A. Whitmer, Paola Gilsanz, Kathryn C. Conlon

**Affiliations:** Department of Epidemiology, School of Public Health, University of Michigan, 1415 Washington Heights, Ann Arbor, MI, 48109, USA; Division of Research, Kaiser Permanente Northern California, 4480 Hacienda Drive, Pleasanton, CA, 94588, USA; Department of Public Health Sciences and Neurology, School of Medicine, University of California Davis, One Shields Avenue, Medical Sciences 1C, Davis, CA, 95616, USA

**Keywords:** Environmental exposures, lead, dementia, epidemiology, cohort, industrial pollution, cognition

## Abstract

**Background:** While lead exposure is associated with poorer cognitive performance in children, the association with late life cognition in diverse cohorts is unknown.

**Method:** In two adult cohorts (Kaiser Health Aging and Diverse Life Experiences (KHANDLE, n=1,638), Study of Healthy Aging in African Americans (STAR, n=741)), we assessed residential proximity to lead releasing facilities, measured through the Toxics Release Inventory, for associations with domain-specific cognition, measured using the Spanish and English Neuropsychological Assessment Scales, two years later. Linear regression models were adjusted for age, sex, race/ethnicity (KHANDLE only), income, education, marital status, smoking status, and alcohol consumption. We meta-analyzed across cohorts.

**Result:** The average age at cognitive test was 76.1 years (KHANDLE) and 68.8 years (STAR) and the average distance between residence and lead releasing facility was 8.2 km (KHANDLE) and 3.6 km (STAR). In meta-analysis, for every 5 km closer a residential address was located to a lead releasing facility we observed −0.05 standard deviation lower verbal episodic memory (95% CI: −0.08, −0.02). Living within a 3 km buffer of a lead releasing facility was associated with −0.10 lower semantic memory (95% CI: −0.18, −0.02) and −0.08 lower global cognition (95% CI: –0.14, −0.02).

**Conclusion:** Residential proximity to a lead releasing facility was associated with poorer cognition two years later among adults in two cohorts. Comprehensive understanding of environmental factors is critical for dementia prevention.

## Introduction

Cognitive impairment and decline in later life are increasingly urgent public health concerns, particularly as today’s older adults are living longer.^1^ Impairment in cognitive domains such as episodic memory, semantic memory, and executive function can significantly compromise independence and overall quality of life, contributing to elevated risks of physical disability and hospitalization.^2^ Even subtle declines in cognitive performance strongly predict the subsequent onset of dementia.^3^ In the United States, nearly two-thirds of adults experience some level of cognitive impairment by age 70.^4^ Dementia, and other related subtypes, rank among the most expensive chronic health conditions in the United States, primarily due to the extended disease duration and the intensive care required.^4,5^ The annual estimated cost of dementia care per individual in 2025 for the U.S., including medical and long term care, was $231.7 billion.^6^ With disease-modifying treatments still limited, identifying modifiable risk factors is a critical strategy for delaying or preventing the onset and progression of dementia.^6^

Environmental toxicants such as air pollution have been identified by the Lancet Commission as a modifiable risk factor for cognitive aging.^6,7^ Another environmental toxicant is lead (chemical symbol Pb). Historically, lead was widely used in gasoline, household paints, tobacco products, water systems, and industrial processes, resulting in extensive environmental contamination and widespread population exposure.^8,9^ Although regulatory measures have reduced average lead levels, legacy contamination persists due to aging infrastructure, industrial emissions, and soil contamination near smelting or recycling facilities.^1,3,9^ Additionally, lead exposure from residentially located lead-releasing industrial facilities are still common in the United States. In 2023 there were 7,507 lead-releasing facilities in the United States, and 425 lead releasing facilities in California.^10^ Many of these facilities are located within significantly populated areas, with some facilities located in counties of more than 100,000 people.^10^ Substantial evidence has linked early life and occupational lead exposures to adverse neurocognitive outcomes, including reduced intellectual functioning and long-term cognitive deficits.^1,11^ The long biological half-life (25-30 years) of lead and preferential accumulation in bone tissue can result in sustained internal exposure that may continue to affect health years after initial contact.^12,13^ Emerging studies suggest that cumulative lead exposure across the life course, even at low-levels, may affect memory, attention, and executive function.^14,15^ However, results are inconsistent and data remain sparse.^9,16^

Little is known about how spatially patterned industrial exposures, such as residential proximity to lead-emitting facilities, relate to cognitive aging in diverse populations. This limitation constrains the ability to evaluate long-term effects of environmental lead exposure in community-based settings, especially in the context of environmental health disparities. Communities with higher proportions of historically racially marginalized populations often experience disproportionate exposure to lead.^17–19^ However, comparatively less attention has been given to evaluating these disparities in lead exposure among adult populations. Few U.S. based studies have evaluated associations between spatial indicators of lead exposure and cognitive outcomes while incorporating lag periods. This is particularly salient in California, where legacy industrial activity and demographic diversity create unique environmental and social exposure contexts. This study addresses these gaps by examining whether residential proximity to lead-releasing facilities is associated with cognitive performance among older adults in California.

Given the limited number of studies, and inconsistent results on industrial lead exposure and cognition in adulthood, we were motivated to perform a geospatial analysis of residential lead exposure across multiple cognitive domains. In this study, we examined whether residential proximity to lead-releasing facilities was associated with verbal episodic memory, semantic memory, executive function, and global cognitive performance, measured two-years later among a diverse group of older adults in California.

## Methods

### Study Participants

This analysis included two mutually exclusive longitudinal cohort studies on aging among older adults in Northern California: the Kaiser Healthy Aging and Diverse Life Experiences (KHANDLE) and the Study of Healthy Aging in African Americans (STAR). The institutional review board at Kaiser Permanente Northern California approved both cohort studies (1278966, 1279068). All participants provided written informed consent.

The KHANDLE cohort included adults ages 65 years and older living in the Sacramento and San Francisco Bay communities of California, USA. To recruit a diverse and representative cohort, stratified random sampling by race, ethnicity, and education was performed.^20^ Participants were eligible for KHANDLE if they were members of Kaiser Permanente Northern California, an integrated healthcare delivery system, and completed at least one voluntary health exam called the Multiphasic Health Checkup (MHC) between the years 1964 and 1985, spoke English or Spanish, and were 65 years or older as of January 1, 2017.^21,22^ Participants were excluded if they had a recorded diagnosis of dementia, chronic obstructive pulmonary disease, congestive heart failure hospitalizations, end-stage renal disease or dialysis.^22–24^ The KHANDLE study is ongoing, and these analyses include information collected at study enrollment, which occurred between April 2017 and December 2018. Further details on cohort design are available.^20,22^ Participants were excluded from the analytic sample if they were missing any cognitive assessment measure (n = 25). To protect participant anonymity due to small sample size, those identifying as Native American were excluded from this analysis.

The STAR cohort included Black adults ages 50 years and older with residential addresses in the San Francisco Bay region of California, USA. Cohort recruitment was performed using stratified random sampling by age and educational attainment.^25^ Participants were eligible for STAR if they were members of Kaiser Permanente Northern California, had at least one MHC visit between 1964 and 1985, identified as African American or Black, and were ages 50 or older on January 1, 2018. Cohort comorbidity exclusion criteria is the same as KHANDLE. The STAR study is ongoing, and these analyses include information collected at study enrollment, which took place between November 2017 and March 2020. Further details on cohort design are available.^25^ Participants were excluded from the analytic sample if they were missing any cognitive assessment measure (n = 5). To protect participant anonymity due to small sample size, those identifying as Native American were excluded from this analysis.

### Study design

In this analysis, each participant had a single exposure measure and a single cognitive outcome measure that were separated by two years in a lagged design (**Figure 1**). Based on the year of a given participant’s cognitive assessment (study range: 2017-2020), we counted backward two years for that participant’s exposure measure (study range: 2015-2018). For example, if a participant had a cognitive test in 2020, we linked their exposure measure from 2018 for analysis.

**Figure 1.**
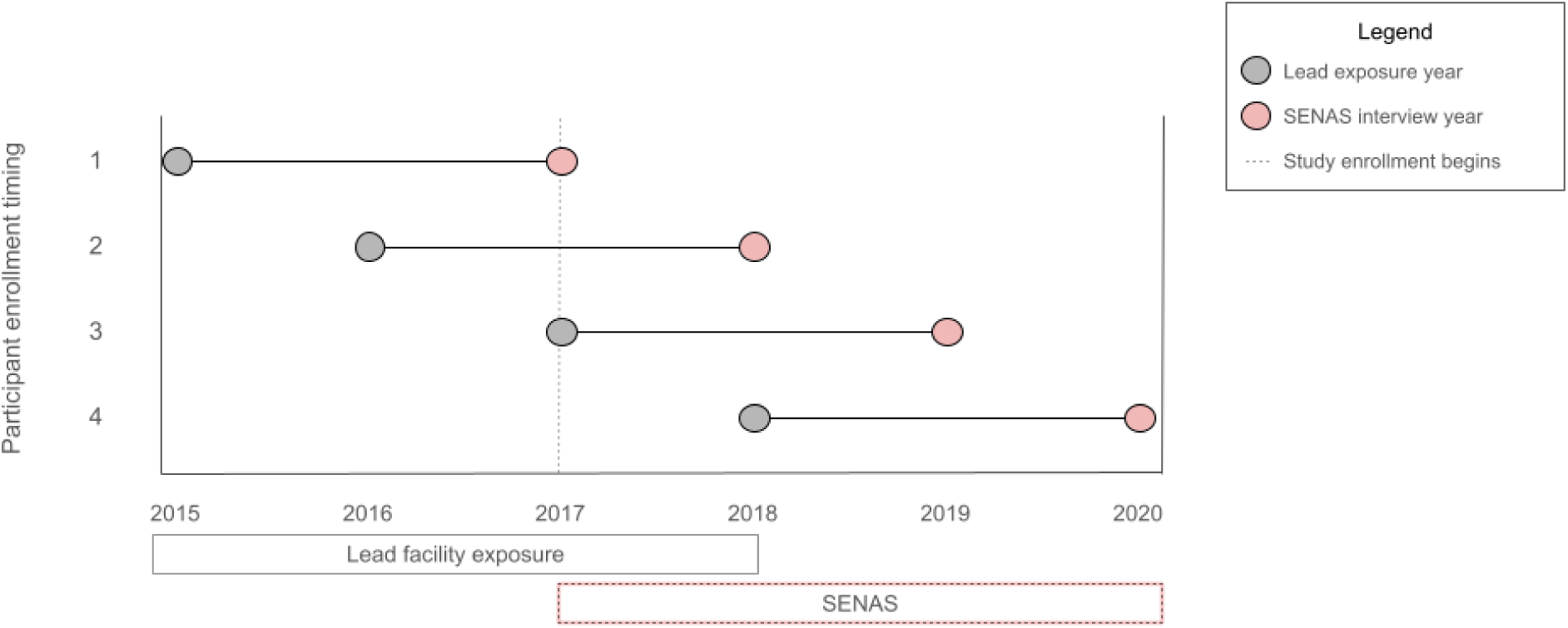
Study design timing: Each participant had a single lead exposure measure (grey) and two years later, had a single cognitive measure (red). The year of each participant’s measures depended on the timing of their cognitive testing, performed using the Spanish and English neuropsychological assessment scales (SENAS). SENAS for all study participants were completed once during enrollment between 2017 and 2020. Thus, the exposure measures generated from residential address proximity to lead releasing facilities were linked between 2015 and 2018. All linkages follow a two year lag design where exposures were measured before cognitive outcomes.

### Exposure measures

The United States Environmental Protection Agency (US EPA) maintains the Toxics Release Inventory (TRI). The TRI, established in 1986, provides open access to nationwide toxic chemical release data reported yearly by industrial and federal facilities across the United States.^26^ The TRI Toxics Tracker stores data from the past 10 years, with detailed information on geography, facility, industry sector, chemical, and release type.^10^ The TRI reports pounds of emissions released from TRI facilities in the following categories: total, air, water, land, off-site, and waste managed. Chemical and facility data for the entire state of California from 2012-2021 were extracted from the TRI Toxics Tracker. To ensure our exposure occurred before our outcome measures, we first restricted the TRI exposure data to the years 2015-2018 (n = 544 facilities). Next, we restricted to facilities releasing lead (designated as either “lead compounds” or “lead”; n = 525 facilities), then to facilities who released more than zero pounds of lead per year (n = 453 facilities).

Residential addresses at ages 40 and 65 were self-reported to each of the cohorts, and we selected the residential address at the age nearest to the study enrollment age for linkage. Participant addresses were geocoded into coordinates of latitude and longitude using ArcGIS (average match score: 98.9 KHANDLE, 97.5 STAR). Geocoded residential addresses were linked to the nearest lead-releasing TRI facility, two years prior to cognitive testing. We created four distance exposure measures which were used in this analysis. We calculated the distance in kilometers from a participant’s residence to the TRI facility, which we used as our continuous exposure variable. To examine buffer sizes around the facilities, we then created three binary variables for living (yes, no) within each of three distances (1.5 km, 3 km, or 5 km) around a lead-releasing TRI facility. The reference group for each categorical distance measure was “no”. We used both continuous and categorical (1.5 km, 3 km, 5 km) exposure measures for statistical analysis.

### Cognitive measures

Cognitive outcomes of verbal episodic memory, semantic memory, executive function, and global cognition were derived from Spanish and English Neuropsychological Assessment Scales (SENAS).^27^ SENAS methodology has been previously described.^27^ Briefly, SENAS was developed with item response theory methodology to allow for valid comparisons of cognition and cognitive change across diverse groups.^27^ The episodic memory domain was a composite measure of item response, category fluency, working memory, learning trials, and delayed free recall.^28^ Semantic memory domain was a composite of object naming, and picture association.^28^ The executive function domain was a composite measure of category fluency, phonemic fluency, and working memory.^28^ Finally, the global cognition domain was an average of the episodic memory, semantic memory, and executive function scores.^27–29^ Descriptions of test administration, development, and guidelines have been described previously.^27^ Each participant completed the SENAS examination once, in either English or Spanish, during baseline interviews between 2017-2020. Interviews were conducted in-person, by study personnel, from 2017-2019. In 2020, interviews were conducted via telephone, due to the COVID-19 pandemic.

Each cognitive outcome was z-score standardized, separately for KHANDLE and STAR, to allow for more meaningful interpretation and comparability across participants. To generally understand cognitive trends within each exposure buffer, we dichotomized each cognitive domain score into high (>0) and low (<=0) for descriptive analyses. Continuous cognitive domain scores were used in statistical analysis.

### Covariate measures

Demographic and health information, including age, sex (male, female), race and ethnicity (Asian, Black, LatinX, Native American, White, or other racial/ethnic identity), education (less than or equal to high school, greater than high school), marital status (married/living as married, not married), smoking status (never, former, current), and alcohol consumption (never, less than once a week, 1-6 days per week, everyday), were collected via in person interview at enrollment. Average household census tract income was acquired from the U.S. Census Bureau database for years 2015-2017, and linked to geocoded residential addresses. For the small number of people who completed study interviews in 2020, we used 2017 average income data for household census tracts, as 2018 data were unavailable.

### Statistical analysis

We performed a single imputation for those missing covariate information on education (n=1, 0.06% KHANDLE; n=3, 0.4% STAR), marital status (n=26, 1.5% KHANDLE; n=17, 2.2% STAR), average income (n=3, 0.1% KHANDLE), smoking status (n=6; 0.4% KHANDLE), and alcohol consumption (n=12; 0.7% KHANDLE) using the Fully Conditional Specification method.^30^ For KHANDLE, race/ethnicity was included as a covariate to inform the imputation. Descriptive statistics were calculated on the included and excluded samples using mean and standard deviation for continuous variables, and frequency and percent for categorical variables. Because KHANDLE and STAR have different geographic, age, and race/ethnicity recruitment strategies, we performed all analyses separately in each cohort. We calculated bivariate sample characteristics by exposure distance (within 1.5 km, 3 km, and 5 km) and high/low values for each cognitive domain (episodic memory, semantic memory, executive function, global cognition), and compared the distributions using Satterthwaite t-test for continuous variables and Fisher’s exact test for categorical variables.

Our primary analyses tested the adjusted associations between residential distance to a lead releasing facility and cognitive score two years later using multivariable linear regression. We evaluated distance to the nearest lead releasing facility as a continuous measure as well as categorically (buffers with radii of 1.5 km, 3 km, 5 km). We examined each distance measure for association with each cognitive domain score, adjusting for age at cognitive testing and sex (minimally adjusted), and further adjusted for race/ethnicity, income, education, marital status, smoking status, and alcohol consumption (fully adjusted). Race and ethnicity was included in fully adjusted models for KHANDLE only, race and ethnicity was not adjusted for in STAR as all participants recruited identified as Black. We performed a conversion of the continuous distance measure, which was modeled as per 1 km increase, by multiplying effect estimates by -5. This allowed us to interpret our findings as per every 5km closer to a lead releasing facility. To increase statistical power and generalizability, we meta-analyzed results across the KHANDLE and STAR cohorts, using random-effects models. For comparison, from these same models, we also reported the associations for a one year increase in age.

### Sensitivity analyses

We conducted multiple sensitivity analyses to interrogate the robustness of our results. We performed a complete case sensitivity analysis to assess how our findings are impacted by different assumptions of missing data. Participants with missing information for any aforementioned covariates were excluded, resulting in final analytic samples of n = 1,590 for KHANDLE, and n = 721 for STAR. Effect modification by age (using categories of <70, 70-74, 75-79,≥ 80) and education (≤ high school, trade school or college, graduate school) were assessed in stratified analyses.

We used SAS statistical software (version 9.4) and R statistical software (version 4.1) for our analyses. Code to produce the analyses is available (https://github.com/bakulskilab).

## Results

### Lead releasing facilities descriptive statistics

The number of lead releasing facilities per county fluctuated by year **(Figure 2)**. Across all exposure years, Los Angeles consistently had the greatest number of TRI facilities per county, with 106 facilities in 2018. The amount of lead released varied by year, mode of release, county, and facility.

**Figure 2.**
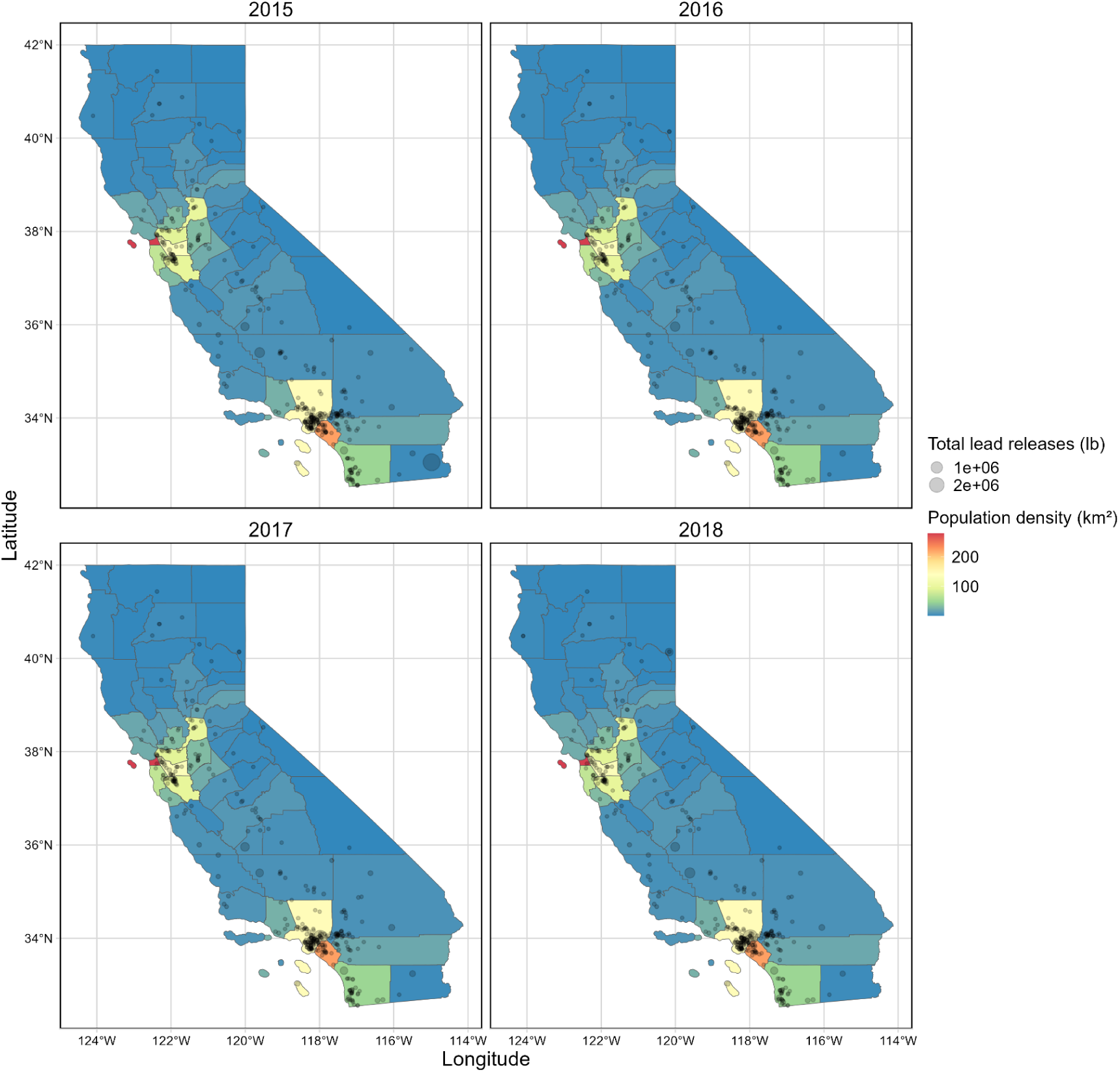
Distribution of Toxics Release Inventory (TRI) lead-releasing facilities by population density in California, years 2015-2018. Points represent the TRI facility location, scaled by the total amount of lead released in pounds per year. California counties are colored by population density per km^2^.

### Study sample descriptive statistics

For the KHANDLE cohort, n = 1,663 participants were eligible for this study. We excluded those without cognitive assessments (n = 25), leaving n = 1,638 included in our analytic sample **(Supplemental Figure 1A)**. Relative to the included sample, those excluded were older (mean 80 vs 76 years), had greater than high school degree (72% vs 83%), lived closer to a TRI facility (mean 7.4 km vs 8.2 km), and were more likely to be Black (36% vs 25%) **(Supplemental Table 1)**. For the STAR cohort, n = 746 participants were eligible for this study. After excluding those without cognitive assessments (n = 5), our included analytic sample size was n = 741 **(Supplemental Figure 1B)**. Relative to the included sample, those excluded were older (mean 69 vs 68 years), had greater than high school degree (80% vs 81.8%), and lived closer to a TRI facility (mean 3 km vs 3.6 km) **(Supplemental Table 1)**.

Among the KHANDLE included sample, the average age at cognitive assessment was 76 years, 58.9% were female, 25.8% identified as Black, 25% as Asian, 19.7% as LatinX, and 29.5% as White **(Table 1)**. The average distance between residence and lead releasing facility was 8.21 km. Among the STAR included sample, the average age at cognitive assessment was 68 years, 68.6% were female, 98.7% identified as Black, and 0.8% LatinX **(Table 2)**. The average distance between residence and lead releasing facility was 3.56 km. In KHANDLE, 60 (3.7%) participants lived within 1.5 km of a lead releasing facility, 326 (19.9%) lived within 3 km of a lead releasing facility, and 742 (45.3%) lived within 5 km. In STAR, 84 (11.3%) participants lived within 1.5 km of a lead releasing facility, 343 (46.3%) lived within 3 km of a lead releasing facility, and 620 (83.7%) lived within 5 km. Overall, those in the STAR cohort tended to live closer to lead releasing facilities, were more likely to be female, and were younger on average, than those in KHANDLE.

**Table 1.**
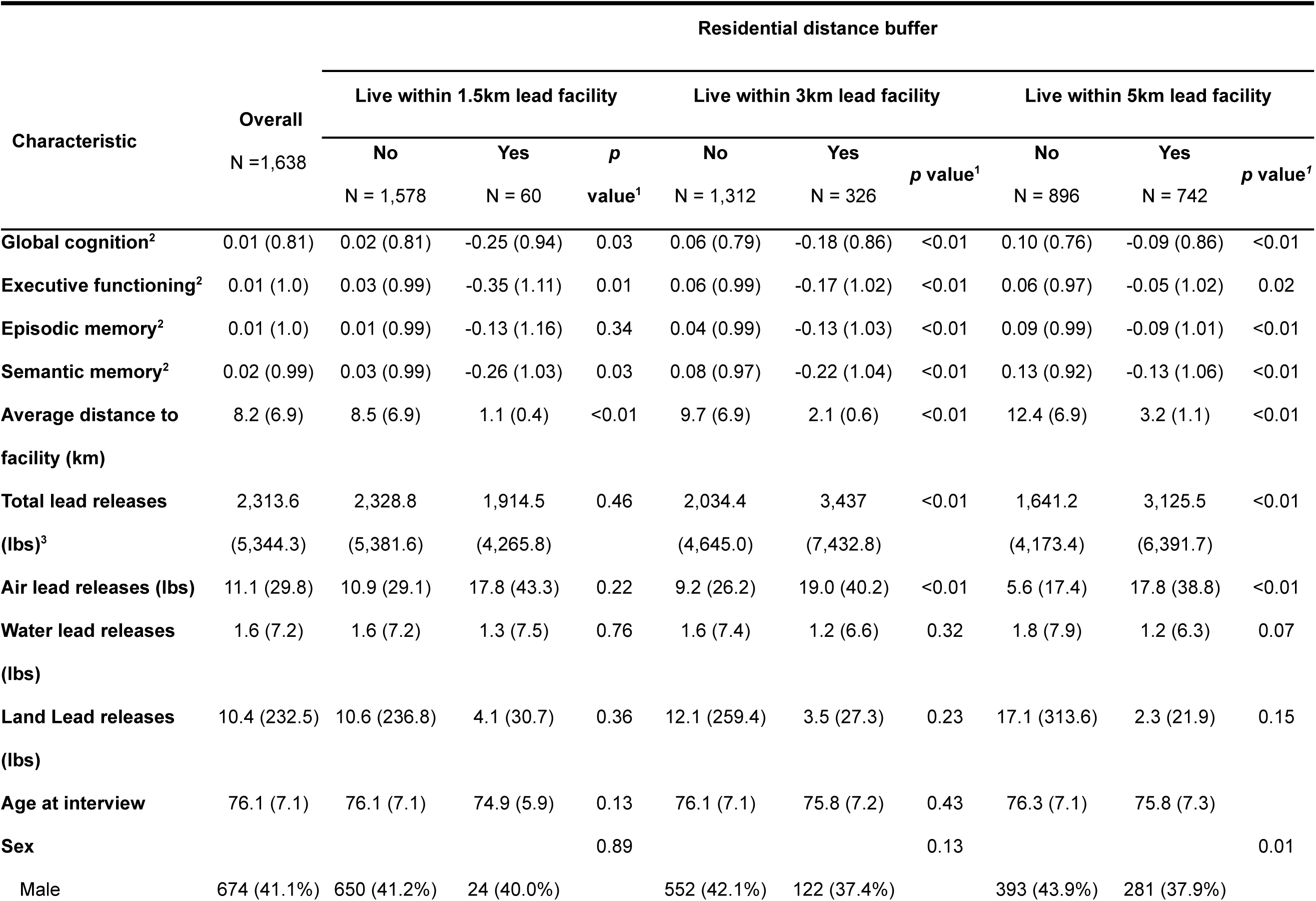
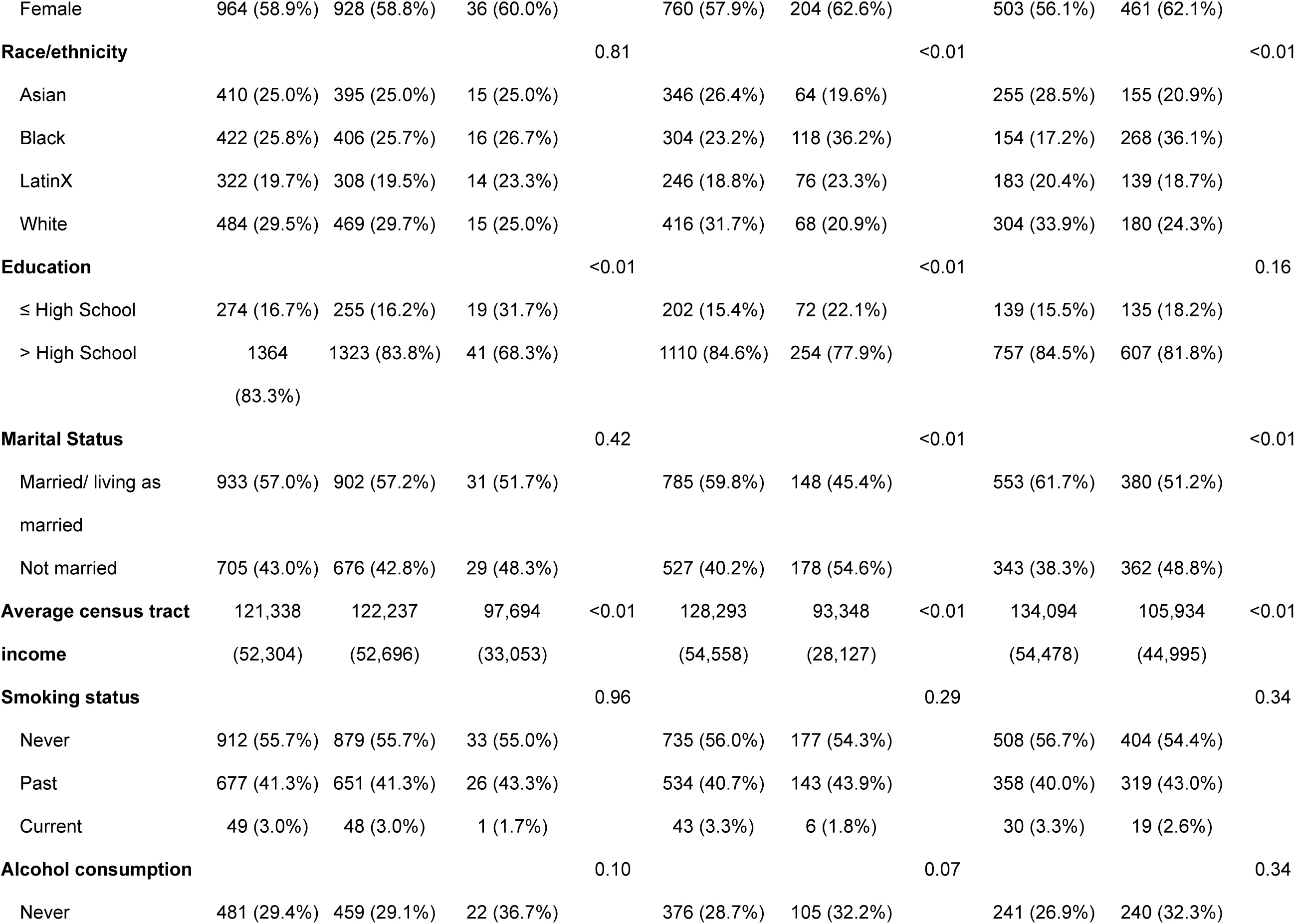
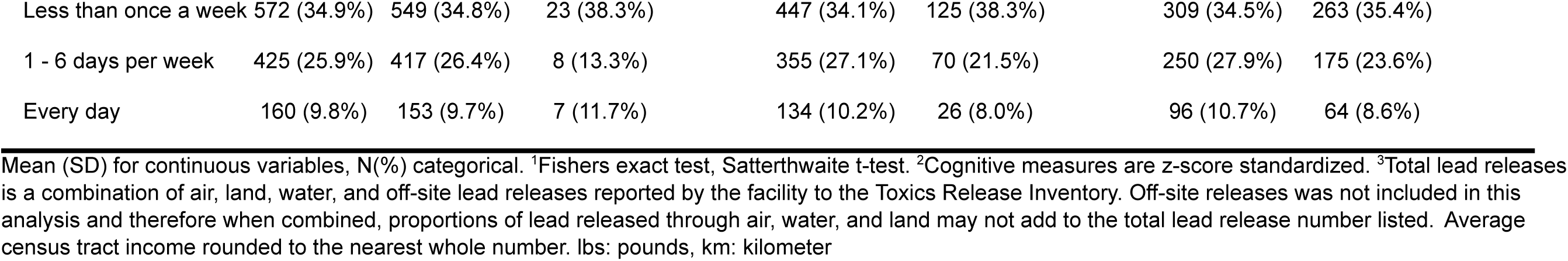
Kaiser Healthy Aging and Diverse Life Experiences Study (KHANDLE) analytic sample descriptive statistics by residential distance buffer around a lead releasing facility.

**Table 2.**
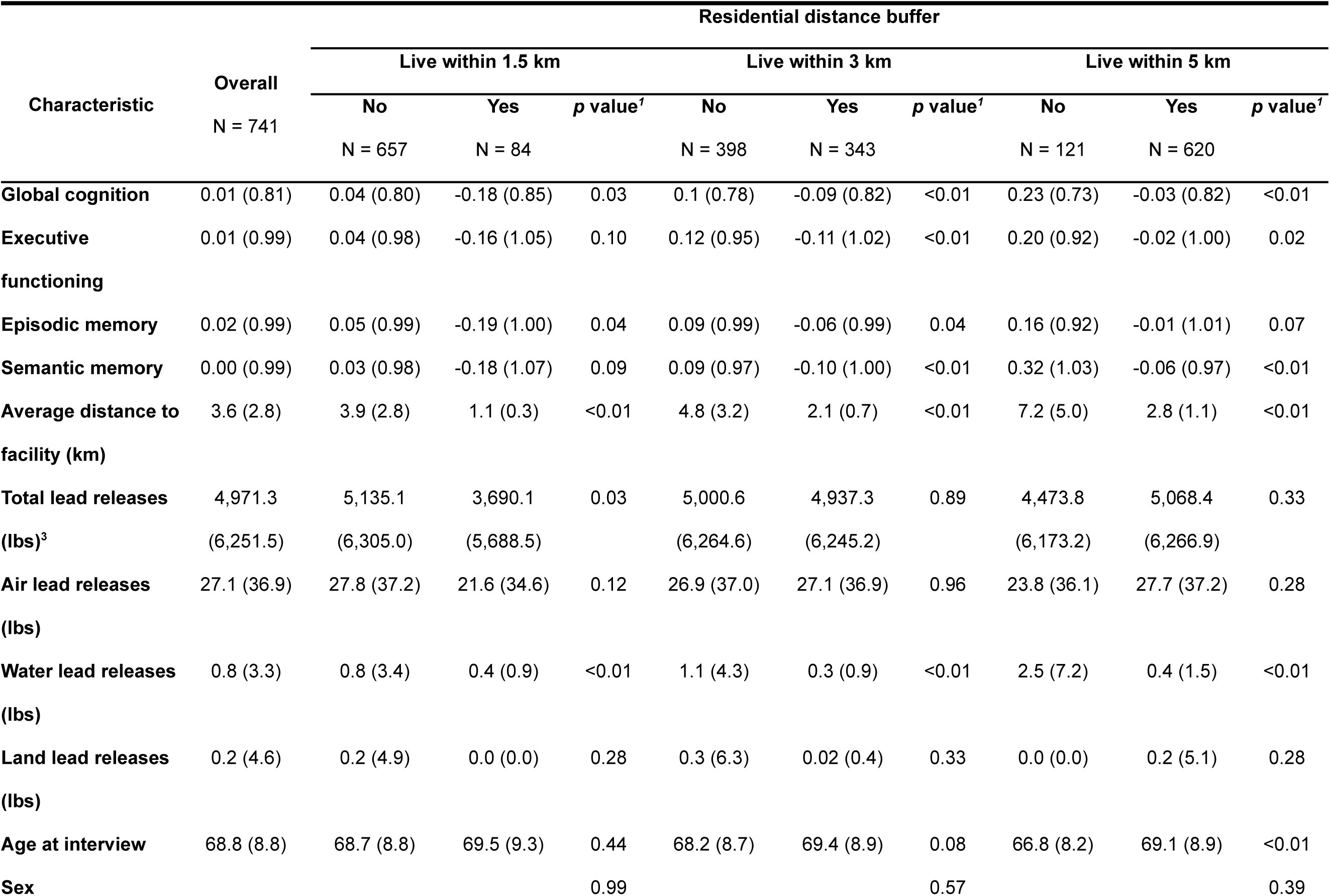
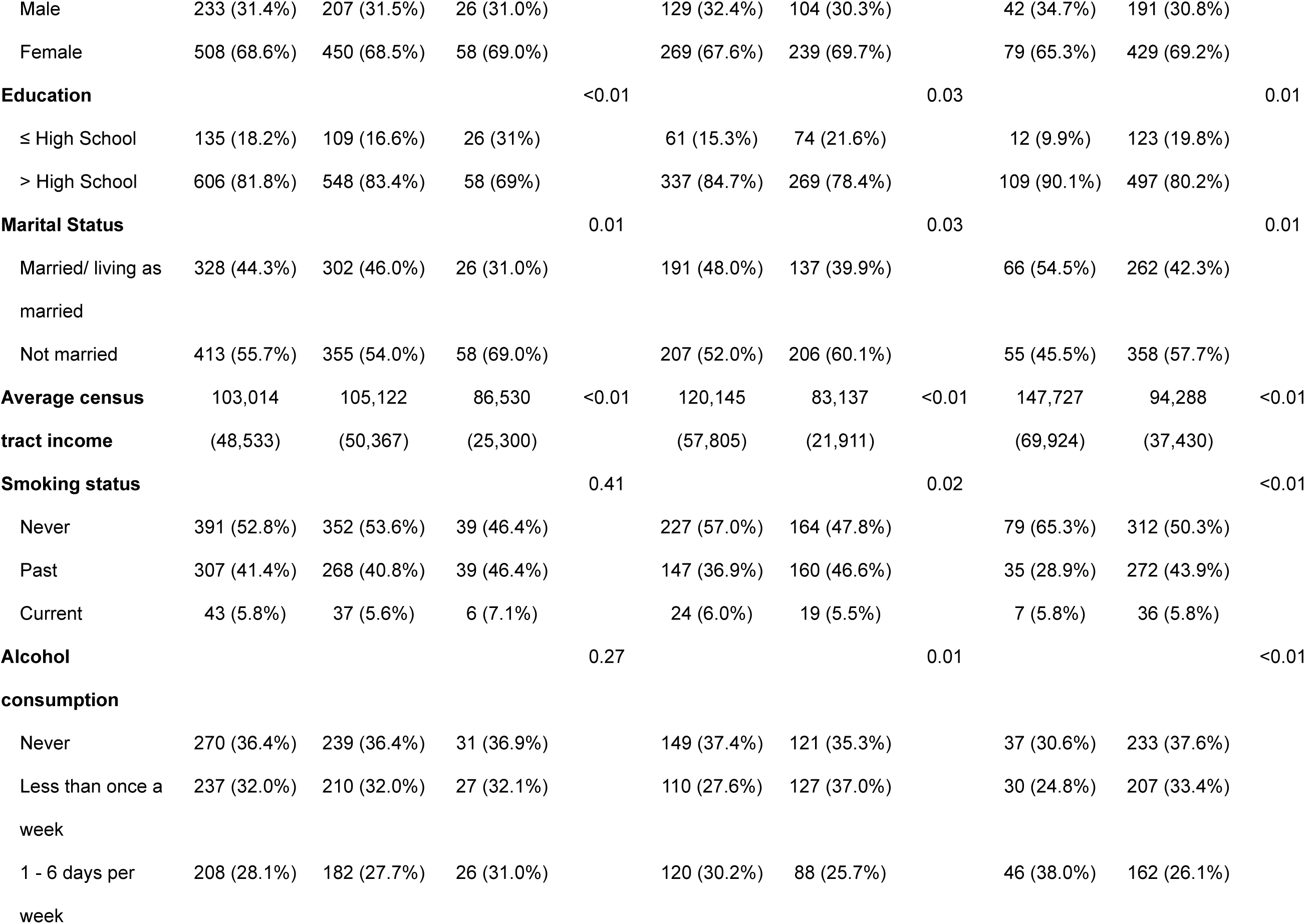
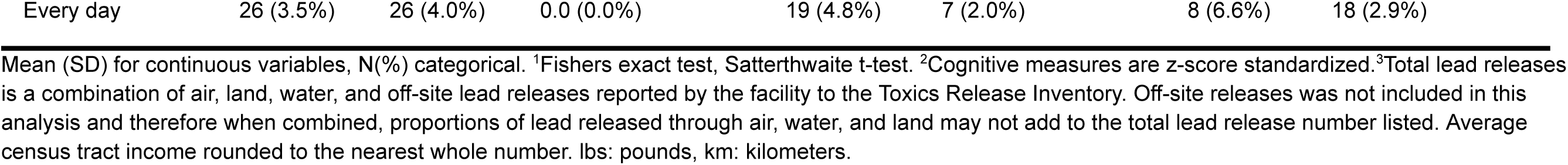
Study of Healthy Aging in African Americans (STAR) analytic sample descriptive statistics by residential distance buffer around a lead releasing facility.

### Residential distance to lead releasing facility association with cognitive domains

When examining bivariate associations with cognitive domain outcomes, participants living within any of the buffers (1.5 km, 3 km, 5 km) were more likely to have lower episodic memory, semantic memory, executive function, and global cognition scores **(Supplemental Tables 2 and 3)**. In KHANDLE fully adjusted models, for every 5 km closer a residential address was located to a lead releasing facility, episodic memory scores two years later were −0.05 standard deviation lower (95% CI: −0.02, −0.08), (**Figure 3 and Supplemental Table 4**). Living within 1.5 km of a lead releasing facility was associated with −0.24 standard deviation lower (95% CI: −0.45, −0.04) executive function scores, two years later. Living within 5 km of a lead releasing facility was associated with −0.15 standard deviation lower (95% CI: −0.24, −0.06) episodic memory scores, and −0.07 standard deviation lower (95% CI: −0.14, −0.01) global cognition scores, two years later. Across categorical exposure buffers, as buffer size increased, we observed slight attenuation of effect estimate. Semantic memory was not associated with continuous or categorical distance measures. Minimally adjusted models were generally stronger than the fully adjusted models **(Supplemental Figure 3 & Supplemental Table 4).**

**Figure 3.**
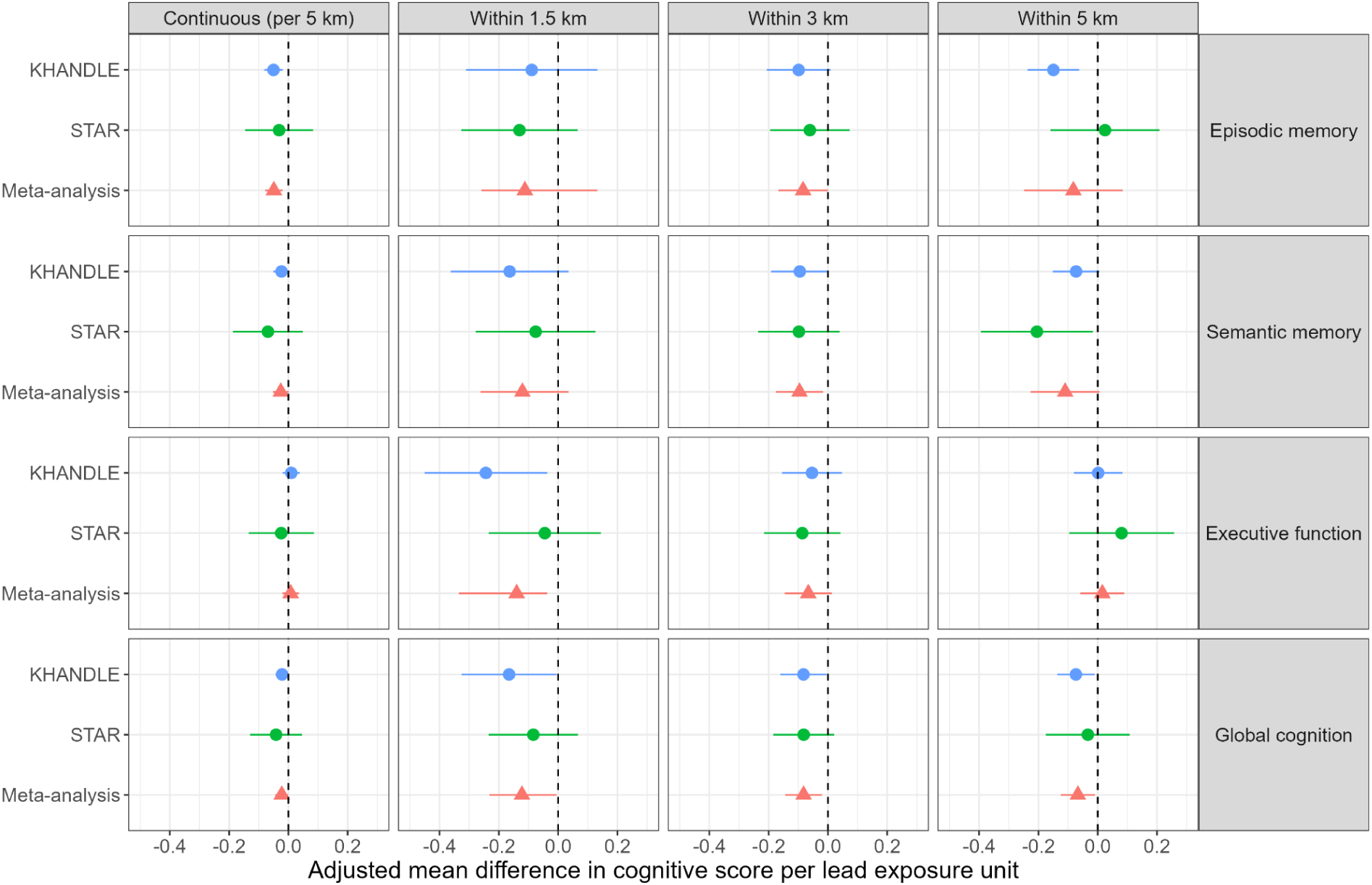
Forest plot of adjusted mean difference cognitive score and 95% confidence intervals by lead measure for KHANDLE (blue) and STAR (green) analytic samples, and meta-analyzed (red) across cohorts. Fully adjusted for age, sex, education, income, marital status, race/ethnicity, smoking status, alcohol consumption. KHANDLE: Kaiser Healthy Aging and Diverse Life Experiences Study; STAR: Study of Healthy Aging in African Americans

In the STAR cohort, in fully adjusted models, living within 5 km of a lead releasing facility was associated with −0.20 standard deviation lower (95% CI: −0.39, −0.02) semantic memory scores, two years later **(Figure 3 and Supplemental Table 5)**. Episodic memory, executive function, and global cognition were not associated with either continuous or categorical distance exposures in fully adjusted models. Again, minimally adjusted models were generally stronger than fully adjusted models **(Supplemental Figure 3 & Supplemental Table 5)**.

Combined meta-analysis estimates showed similar magnitude and direction of effect with cohort specific main models **(Supplemental Table 6)**. In fully adjusted meta-analysis models, for every 5 km closer a residence was located to a lead releasing facility, episodic memory scores two years later were −0.05 standard deviation lower (95% CI: −0.08, −0.02).

For comparison, in meta-analysis, a one-year increase in age was associated with −0.04 standard deviation lower episodic memory (95% CI: −0.05, −0.04), −0.04 standard deviation lower semantic memory (95% CI: −0.04, −0.04), −0.05 standard deviation lower executive function (95% CI: −0.05, −0.04), and −0.04 standard deviation lower global cognition (95% CI: −0.05, −0.04).

### Sensitivity analysis

Complete case descriptive statistics are available **(Supplemental Table 7)**. Overall, we observed associations with consistent magnitude with slight attenuation, compared to main models **(Supplementary Tables 8 and 9)**. In KHANDLE fully adjusted models, living within the 5 km exposure buffer, was associated with −0.13 standard deviation lower (95% CI: −0.22, −0.05) episodic memory scores and −0.07 standard deviation lower (95% CI: −0.13, −0.002) global cognition scores, two years later. In STAR fully adjusted models, lower cognition was generally observed, but this only reached statistical significance with 5km buffer exposure to a TRI facility and semantic memory (β:-0.19, 95% CI: −0.38, −0.004).

In age stratified models, the associations between residential distance to a lead releasing facility and lower cognitive domain scores remained consistent with main models **(Supplementary Tables 10 and 11)**. The largest magnitudes of association were observed for participants in the 70-74 and 75-79 age categories. In KHANDLE, for ages 70-74, living within 5 km of a lead releasing facility was associated with −0.23 standard deviation (95% CI: −0.39, −0.08) lower episodic memory scores, two years later. In STAR, for ages 75-79, living within 5 km of a lead releasing facility was associated with −0.54 standard deviation (95% CI: -1.05, −0.03) lower episodic memory scores, two years later.

For education stratified models, the magnitude and direction of associations were consistent with the main models **(Supplementary Tables 12 and 13)**. The largest magnitudes were observed for those reporting an education level of less than or equal to high school. In KHANDLE fully adjusted models among those with less than or equal to a high school education, living within the 5 km exposure buffer, lower cognitive scores were observed for episodic memory (β:-0.29, 95% CI: −0.51, −0.07), semantic memory (β:-0.36, 95% CI: −0.56, −0.15), and global cognition (β:-0.26, 95% CI: −0.43, −0.10). Among those with higher educational attainment, lead exposure was also associated with lower cognitive scores. In KHANDLE among those with a graduate degree, living within the 3km buffer was associated with −0.27 standard deviation (95% CI: −0.50, −0.05) lower semantic memory scores, two years later, in fully adjusted models. In the STAR cohort, a similar trend was observed.

## Discussion

Overall, in two California longitudinal cohorts on aging with harmonized exposure and outcome measures, we found that closer residential distance to lead releasing facilities was associated with worse cognitive scores two years later. We examined the association between residential distance to a lead-releasing facility as a continuous measure as well as categorically (buffers with radii of 1.5 km, 3 km, 5 km) and domain specific cognition two years later. Our most consistent finding, across cohorts and sensitivity analyses, was an association between residing within 5 km of a lead releasing facility and lower cognitive scores. Specifically, in the meta-analysis, living within 5 km of a lead releasing facility was associated with −0.07 standard deviation (95% CI: −0.12, −0.0009) lower global cognition scores two years later. The direction of effect was consistent when we tested distance on a continuous scale and examined buffers of 1.5 km and 3 km. This study suggests that residential lead exposure may be associated with lower cognitive performance, and could be a potential modifiable risk factor for dementia.

Our findings align with work from the Cardiovascular Health Cognition Study (residences in Sacramento County, CA; Washington County, MD; Forsyth County, NC; Pittsburgh, PA) demonstrating an association between industrial chemical releases and dementia risk in adults.^15^ That study used an area model of cumulative TRI across chemicals weighted by their estimated toxicity and incorporating mode of release called the Risk-Screening Environmental Indicators (RSEI) model, created by the EPA. Higher RSEI scores relate to higher concentrations of hazardous chemicals released.^31^ The Cardiovascular Health Cognition Study observed each RSEI score doubling was associated with 1.09 times higher odds of prevalent dementia (95%CI: 1.00, 1.19) and 1.16 times higher hazard of incident vascular dementia (95%CI: 1.01, 1.34).^15^ Past studies in children are also consistent with our findings, reporting higher toxic air emissions estimated by RSEI were associated with worse cognitive performance.^32–34^ Other adverse health impacts have also been associated with nearby industrial facilities releasing toxic chemicals.^32–36^ These prior works on residential proximity to industrial releases integrated across chemicals using RSEI used the same underlying TRI data source as this study. We chose to study lead specifically based on our previous research, our *a priori* hypothesis about lead toxicity on dementia related pathways, and to improve the potential translation to policy intervention from our findings.^37,38^

Most existing lead and dementia research has implemented biomarker-based exposure assessments, such as blood lead levels, which primarily reflect recent exposure and may not capture chronic accumulation with environmental contamination.^36,39^ Furthermore, little is known about how spatially patterned industrial exposures, such as residential proximity to lead-emitting facilities, relates to cognitive aging in diverse populations. This limitation constrains the ability to evaluate long-term effects of environmental lead exposure in community-based settings, especially in the context of environmental health disparities. Communities with higher proportions of historically racially marginalized populations often experience disproportionate exposure to lead, which has been well documented particularly among children.^18,19^ However, comparatively less attention has been given to evaluating these disparities in lead exposure among adult populations. Few US-based studies have evaluated associations between spatial indicators of lead exposure and cognitive outcomes. This is particularly salient in California, where legacy industrial activity and demographic diversity create unique environmental and social exposure contexts. This study addressed these gaps by examining whether residential proximity to lead-releasing facilities was associated with memory performance among older adults in California.

In the present analysis, closer residential distances to lead releasing facilities were associated with lower domain-specific cognitive performance two years later. These findings may indicate that exposures in adulthood are related to cognition, and potentially contribute to dementia development. Our results differed by exposure distance metric and cognitive domain, with episodic memory having the strongest associations across exposure distances. Potentially, different cognitive domains may be more influenced by exposure to lead. We also observed lower cognitive scores among those with lower education, which is consistent with previous literature.^32,40,41^ However, for the relationship between lead and cognition, we did not observe effect modification by education. These results indicate that more education is not sufficient to mitigate the relationship between lead and cognition, as those with higher education were also observed to have an association between lead and lower cognition. Our findings highlight that residential lead exposure in adulthood is not only an environmental health concern, but also an environmental justice concern. To the best of our knowledge, this is the first study using TRI facility data to assess associations with domain specific cognition in adults.

Lead is a well-established neurotoxicant that can cross the blood–brain barrier and accumulate in brain tissue.^42^ Once in the central nervous system, it disrupts neurophysiological process through multiple pathways, including interfering with calcium-mediated signaling and neurotransmission, promoting oxidative stress, altering membrane fluidity and permeability, and mimicking calcium ions (Ca²⁺) to impair intracellular signaling and synaptic activity.^42–45^ These biological disruptions, combined with emerging evidence of lead’s role in inducing neuroinflammation, create a pathological environment that may accelerate cognitive decline in aging populations. Lead exposure is also associated with numerous related adverse health outcomes, such as hypertension, a documented modifiable risk factor for dementia.^6,37^ The impact of lead on cognitive function may occur through both the direct neurotoxic mechanisms and indirect pathways.^38,39,42^ For example, lead-induced hypertension and cardiovascular dysfunction have been linked to cognitive impairment and dementia.^39,43^ Biological evidence strongly supports a mechanistic link between lead exposure and neurodegeneration. However, epidemiologic evidence in general older adult populations with environmentally relevant exposures remains limited. Few population-based studies have assessed the cognitive impacts of environmental lead exposure in community-dwelling older adults, particularly using geospatial methods to characterize long-term environmental burden.^15^

### Limitations and future directions

Our analysis has several limitations. The smaller sample size for the STAR cohort, could potentially limit the ability to detect associations between our exposure and outcome measures. Also, distance measures were used as a proxy for exposure, rather than an internal exposure biomarker, which could potentially lead to exposure misclassification. Distance measures may not accurately reflect actual exposure levels, and assumes exposure levels within a distance measure are uniform for all participants. However, distance based models are considered appropriate for studies when the etiology of health effects is unclear, and provides beneficial exploratory analysis to identify directions for more sophisticated future analyses.^35,46–48^. TRI facilities that release lead may release other toxic substances that may be harmful to cognition, so the distance to these lead releasing facilities may represent proximity to other toxic releases as well. Although we focused on TRI facilities that released lead from 2015-2018, living near these facilities could have exposed people to decades of toxic releases prior to those years. Additionally, we used only one time point to assess prior lead exposure, which does not reflect the true life-time lead exposure for each participant. In future analyses, we plan to incorporate quantities of lead releases, additional cognitive assessments and longer lag exposure windows to investigate how cognition trajectories are associated with residential exposure to lead releasing facilities.

### Strengths

A significant strength of this study is the diverse population, which allowed us the opportunity to study exposure effects in largely underrepresented populations who were enrolled in healthcare plans and had equal access to healthcare. In addition, our two-year lagged study design between exposure and cognitive measure is another strength. This model design affords us information on how lead exposure impacts domain specific cognition over relatively short periods of time. Finally, how lead exposure in adulthood contributes to cognitive performance, and dementia risk is understudied, this analysis helps to fill this knowledge gap. Lead is a well-documented early life neurotoxicant, however, pertinent time periods for exposures in adulthood and their contribution to cognition requires investigation, particularly for diverse cohorts. Our findings together with the literature indicate both adult and childhood lead exposures may be important factors to consider when assessing lifetime risk of dementia.

### Summary and implications

In this analysis, closer residential proximity to lead releasing facilities was associated with lower episodic memory, semantic memory, executive function, and global cognition, two years before cognitive testing. We found that living in close proximity to a lead releasing facility may contribute to worse cognitive performance. Future studies incorporating follow-up cognitive assessments and additional exposure years are needed to help understand associations between lead exposure in adulthood and cognitive decline. Lead exposure is still widespread and variable in the US population. Lead has health impacts throughout the life course, and one prevention strategy to maintain cognitive health is to continue to reduce lead exposure levels in the population. Society-wide types of interventions to reduce lead exposure include further policy and regulation changes to reduce the amount of lead that is produced and released commercially, and to abate or clean up existing lead in the environment. Individual-level changes to reduce lead exposure include ensuring adequate iron and calcium intake and if living in housing built prior to 1978, regularly cleaning windowsills and floors, covering chipping paint, and avoiding projects that create lead paint dust.^49^ Policies and individual practices to reduce lead exposure may be effective for preventing cognitive impairment among older adults.

## Supporting information

All supplemental tables and figures

## Sources of funding

The Kaiser Health Aging and Diverse Life Experiences and the Study of Healthy Aging in African Americans are supported by the National Institute on Aging (RF1AG052132, RF1AG050782). This analysis was supported by the National Institute on Aging (R01AG074347 and R01-1AG067525).

## Acknowledgements

We appreciate the participants and staff of the Kaiser Health Aging and Diverse Life Experiences and the Study of Healthy Aging in African Americans.

## Conflicts of interest

Declarations of interest: none

## Data availability

All lead exposure data used for this analysis are publicly available and can be downloaded from the EPA TRI Toxics Tracker webpage at https://edap.epa.gov/public/extensions/TRIToxicsTracker/TRIToxicsTracker.html.

For access to participant data used in this analysis, please contact the senior author, Kathryn Conlon.

## Notes

### Competing Interest Statement

The authors have declared no competing interest.

### Author Declarations

The institutional review board at Kaiser Permanente Northern California approved both cohort studies (1278966, 1279068)

