## Supplementary material for "Association between proximity to a lead releasing facility and cognition in diverse cohorts": All supplemental tables and figures

Supplemental tables: 13; Supplemental figures: 3.

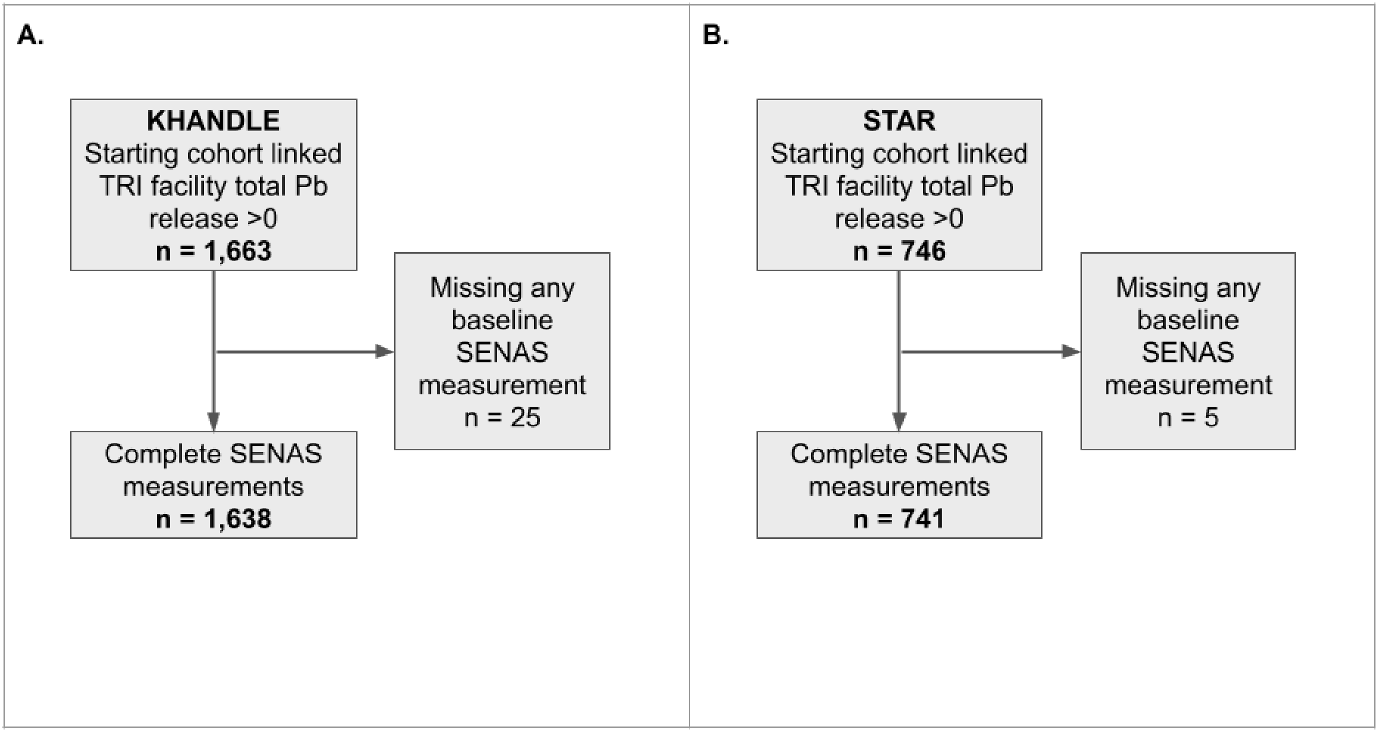


**Supplemental Figure 1**. Flowchart for study inclusion. Panel A represents selection for Kaiser Healthy Aging and Diverse Life Experiences Study (KHANDLE). Panel B represents selection for Study of Healthy Aging in African Americans (STAR). SENAS: Spanish and English neuropsychological assessment scales; TRI: Toxics release inventory; Pb: lead.

**Supplementary Table 1.** Descriptive statistics for the included and excluded participants in the Kaiser Healthy Aging and Diverse Life Experiences Study (KHANDLE) and Study of Healthy Aging in African Americans (STAR) cohorts.

| | Cohort | | | | | | | |
|---|---|---|---|---|---|---|---|---|
| | KHANDLE | | | | STAR | | | |
| **Characteristic** | **Overall** N = 1,663 | **Excluded** N = 25 | **Included** N = 1,638 | ***p* value**[1] | **Overall,** N = 746 | **Excluded,** N = 5 | **Included,** N = 741 | ***p* value**[1] |
| **Global cognition**[2] | 0.01 (0.81) | 0.79 (0.00) | 0.01 (0.81) | 0.07 | 0.01 (0.81) | - | 0.01 (0.81) | 0.02 |
| (missing) | 1 | 1 | - | | | | | |
| **Executive function**[2] | 0.01 (1.00) | -0.28 (0.99) | 0.01 (1.00) | <0.01 | 0.01 (0.99) | -1.15 (1.22) | 0.01 (0.99) | <0.01 |
| (missing) | 10 | 10 | - | | | | | |
| **Episodic memory**[2] | 0.01 (1.00) | -0.05 (0.94) | 0.01 (1.00) | 0.60 | 0.02 (0.99) | -0.46 (0.65) | 0.02 (0.99) | 0.08 |
| (missing) | 4 | 4 | - | | 2 | 2 | - | |
| **Semantic memory**[2] | 0.01 (1.00) | -0.40 (1.39) | 0.02 (0.99) | 0.06 | 0.00 (0.99) | -0.09 (0.42) | 0.00 (0.99) | 0.03 |
| (missing) | 7 | 7 | - | | 3 | 3 | - | |
| **Average distance to facility (km)** | 8.2 (6.9) | 7.4 (6.8) | 8.2 (6.9) | 0.36 | 3.6 (2.8) | 3.0 (1.7) | 3.6 (2.8) | 0.44 |
| **Lead releasing facility within 1.5km** | | | | 0.19 | | | | 0.34 |
| No | 1602 (96.3%) | 24 (96.0%) | 1578 (96.3%) | | 661 (88.6%) | 4 (80.0%) | 657 (88.7%) | |
| Yes | 61 (3.7%) | 1 (4.0%) | 60 (3.7%) | | 85 (11.4%) | 1 (20.0%) | 84 (11.3%) | |
| **Lead releasing facility within 3km** | | | | 0.46 | | | | 0.16 |
| No | 1331 (80.0%) | 19 (76.0%) | 1312 (80.1%) | | 401 (53.8%) | 3 (60.0%) | 398 (53.7%) | |
| Yes | 332 (20.0%) | 6 (24.0%) | 326 (19.9%) | | 345 (46.2%) | 2 (40.0%) | 343 (46.3%) | |
| **Lead releasing facility within 5km** | | | | 0.23 | | | | 0.16 |
| No | 907 (54.5%) | 11 (44.0%) | 896 (54.7%) | | 122 (16.4%) | 1 (20.0%) | 121 (16.3%) | |
| Yes | 756 (45.5%) | 14 (56.0%) | 742 (45.3%) | | 624 (83.6%) | 4 (80.0%) | 620 (83.7%) | |
| **Total lead releases (lbs)**[3] | 2,307.5 (5,331.9) | 1,908.6 (4,516.4) | 2,313.6 (5,344.3) | 0.90 | 4,956.1 (6,245.5) | 2,712.0 (5,351.4) | 4,971.3 (6,251.5) | 0.22 |

| | | | | | | | | |
|---|---|---|---|---|---|---|---|---|
| **Air lead releases (lbs)** | 11.3 (30.1) | 20.5 (47.6) | 11.1 (29.8) | 0.30 | 26.9 (36.9) | 17.1 (38.2) | 27.1 (36.9) | 0.29 |
| **Water lead releases (lbs)** | 1.5 (7.2) | 0.4 (2.2) | 1.6 (7.2) | 0.56 | 0.8 (3.3) | 0.0 | 0.8 (3.3) | <0.01 |
| **Land lead releases (lbs)** | 10.2 (230.7) | 0.0 | 10.4 (232.5) | 0.07 | 0.2 (4.6) | 0.0 | 0.2 (4.6) | 0.28 |
| **Age at interview** | 76.1 (7.2) | 80.3 (8.9) | 76.1 (7.1) | 0.01 | 68.8 (8.8) | 69.5 (5.7) | 68.8 (8.8) | 0.50 |
| **Sex** | | | | 0.90 | | | | 0.19 |
| Male | 684 (41.1%) | 10 (40.0%) | 674 (41.1%) | | 235 (31.5%) | 2 (40.0%) | 233 (31.4%) | |
| Female | 979 (58.9%) | 15 (60.0%) | 964 (58.9%) | | 511 (68.5%) | 3 (60.0%) | 508 (68.6%) | |
| **Race/ethnicity** | | | | <0.01 | | | | <0.01 |
| Asian | 412 (24.8%) | 2 (8.0%) | 410 (25.0%) | | - | - | - | |
| Black | 431 (25.9%) | 9 (36.0%) | 422 (25.8%) | | 736 (98.7%) | 5 (100.0%) | 731 (98.7%) | |
| LatinX | 327 (19.7%) | 5 (20.0%) | 322 (19.7%) | | 6 (0.8%) | - | 6 (0.8%) | |
| Native American | 3 (0.2%) | 3 (12.0%) | - | | 4 (0.5%) | - | 4 (0.5%) | |
| White | 490 (29.5%) | 6 (24.0%) | 484 (29.5%) | | - | - | - | |
| **Education** | | | | <0.01 | | | | <0.01 |
| ≤ High School | 281 (16.9%) | 7 (28.0%) | 274 (16.7%) | | 136 (18.2%) | 1 (20.0%) | 135 (18.2%) | |
| > High School | 1382 (83.1%) | 18 (72.0%) | 1364 (83.3%) | | 610 (81.8%) | 4 (80.0%) | 606 (81.8%) | |
| **Marital Status** | | | | <0.01 | | | | <0.01 |
| Married/ living as married | 944 (56.8%) | 11 (44.0%) | 933 (57.0%) | | 329 (44.1%) | 1 (20.0%) | 328 (44.3%) | |
| Not Married | 714 (42.9%) | 9 (36.0%) | 705 (43.0%) | | 416 (55.8%) | 3 (60.0%) | 413 (55.7%) | |
| (missing) | 5 | 5 | - | | 1 | 1 | - | |
| **Average census tract income** | 1,213,78 (52,401) | 123,999 (59,546) | 121,338 (52,304) | 0.68 | 103,178 (48,673) | 127,476 (688,045) | 103,014 (48,533) | 0.81 |
| **Smoking status** | | | | <0.01 | | | | <0.01 |
| Never | 920 (55.3%) | 8 (32.0%) | 912 (55.7%) | | 396 (53.1%) | 5 (100%) | 391 (52.8%) | |

| | | | | | | | | |
|---|---|---|---|---|---|---|---|---|
| Former | 691 (41.6%) | 14 (56.0%) | 677 (41.3%) | | 307 (41.2%) | - | 307 (41.4%) | |
| Current | 52 (3.1%) | 3 (12.0%) | 49 (3.0%) | | 43 (5.8%) | - | 43 (5.8%) | |
| **Alcohol consumption** | | | | <0.01 | | | | <0.01 |
| Never | 492 (29.6%) | 11 (44.0%) | 481 (29.4%) | | 274 (36.7%) | 4 (80%) | 270 (36.4%) | |
| Less than once a week | 580 (34.9%) | 8 (32.0%) | 572 (34.9%) | | 237 (31.8%) | - | 237 (32.0%) | |
| 1-6 days per week | 430 (25.9%) | 5 (20.0%) | 425 (25.9%) | | 208 (27.9%) | - | 208 (28.1%) | |
| Every day | 161 (9.7%) | 1 (4.0%) | 160 (9.8%) | | 27 (3.6%) | 1 (20%) | 26 (3.5%) | |

Mean (SD) for continuous variables, N(%) categorical. [1]Fishers exact test, Satterthwaite t-test. [2]Cognitive measures are z-score standardized. [3]Total lead releases is a combination of air, land, water, and off-site lead releases reported by the facility to the Toxics Release Inventory. Off-site releases was not included in this analysis and therefore when combined, proportions of lead released through air, water, and land may not add to the total lead release number listed. Average census tract income rounded to the nearest whole number. lbs: pounds, km: kilometers

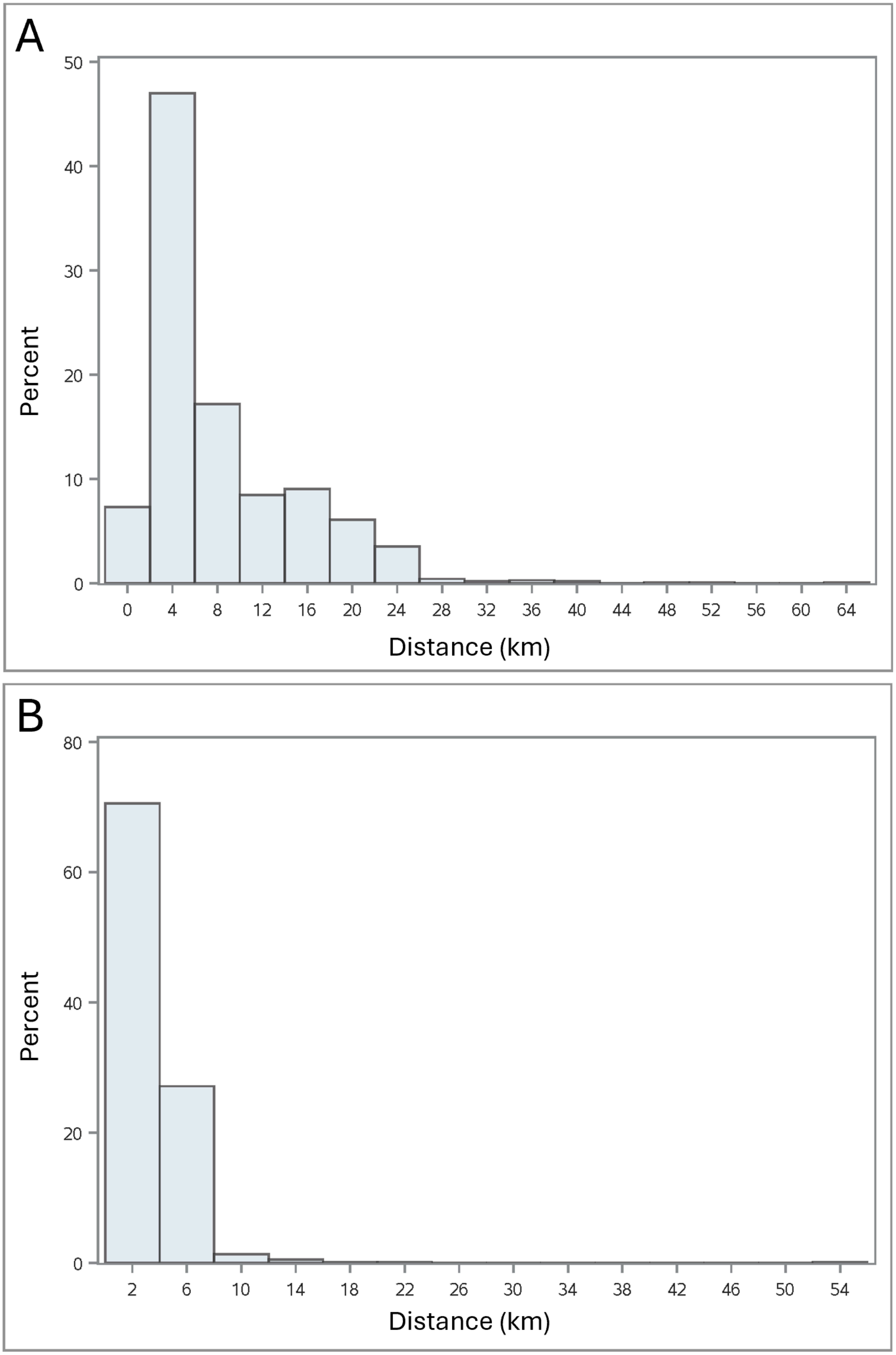


**Supplemental Figure 2.** Distributions of residential distance to lead releasing facility in kilometers (km) in analytic sample by cohort. Panel A represents Kaiser Healthy Aging and Diverse Life Experiences Study (KHANDLE). Panel B represents Study of Healthy Aging in African Americans (STAR). km: kilometers.

**Supplemental Table 2.** Kaiser Healthy Aging and Diverse Life Experiences Study (KHANDLE) analytic descriptive statistics by baseline cognitive domain test results.

| **Characteristic** | **Overall** | **Cognitive domain tested** | | | | | | | | | | | |
|---|---|---|---|---|---|---|---|---|---|---|---|---|---|
| | | **Global cognition** | | | **Executive functioning** | | | **Episodic memory** | | | **Semantic memory** | | |
|  | N = 1,638 | **Low** N = 783 | **High** N = 855 | ***p* value**[1] | **Low** N = 820 | **High** N = 818 | ***p* value**[1] | **Low** N =808 | **High** N = 830 | ***p* value**[1] | **Low** N = 764 | **High** N = 874 | ***p* value**[1] |
| **Average distance to facility (km)** | 8.2 (6.9) | 7.8 (7.1) | 8.6 (6.8) | 0.03 | 8 (6.8) | 8.4 (7) | 0.29 | 7.9 (7.1) | 8.5 (6.7) | 0.05 | 7.6 (6.5) | 8.8 (7.2) | <0.01 |
| **Lead facility within 1.5 km** | | | | 0.18 | | | 0.08 | | | 0.89 | | | 0.06 |
| No | 1578 (96.3%) | 749 (95.7%) | 829 (97.0%) | | 783 (95.5%) | 795 (97.2%) | | 779 (96.4%) | 799 (96.3%) | | 729 (95.4%) | 849 (97.1%) | |
| Yes | 60 (3.7%) | 34 (4.3%) | 26 (3.0%) | | 37 (4.5%) | 23 (2.8%) | | 29 (3.6%) | 31 (3.7%) | | 35 (4.6%) | 25 (2.9%) | |
| **Lead facility within 3 km** | | | | <0.01 | | | <0.01 | | | 0.10 | | | <0.01 |
| No | 1312 (80.1%) | 605 (77.3%) | 707 (82.7%) | | 635 (77.4%) | 677 (82.8%) | | 634 (78.5%) | 678 (81.7%) | | 579 (75.8%) | 733 (83.9%) | |
| Yes | 326 (19.9%) | 178 (22.7%) | 148 (17.3%) | | 185 (22.6%) | 141 (17.2%) | | 174 (21.5%) | 152 (18.3%) | | 185 (24.2%) | 141 (16.1%) | |
| **Lead facility within 5 km** | | | | <0.01 | | | 0.04 | | | 0.01 | | | <0.01 |
| No | 896 (54.7%) | 393 (50.2%) | 503 (58.8%) | | 428 (52.2%) | 468 (57.2%) | | 417 (51.6%) | 479 (57.7%) | | 376 (49.2%) | 520 (59.5%) | |
| Yes | 742 (45.3%) | 390 (49.8%) | 352 (41.2%) | | 392 (47.8%) | 350 (42.8%) | | 391 (48.4%) | 351 (42.3%) | | 388 (50.8%) | 354 (40.5%) | |

| | | | | | | | | | | | | | |
|---|---|---|---|---|---|---|---|---|---|---|---|---|---|
| **Total lead releases (lbs)**[3] | 2,313.6 (5,344.3) | 2,811.7 (6,111.4) | 1,857.5 (4,484.8) | <0.01 | 2,640.8 (5,988.9) | 1,985.6 (4,588.9) | 0.01 | 2,644.4 (5,572.9) | 1,991.6 (5,094.7) | 0.01 | 2,794.9 (6,111.8) | 1,892.8 (4,531.0) | <0.01 |
| **Air lead releases (lbs)** | 11.1 (29.8) | 13.3 (31.9) | 9.1 (27.5) | <0.01 | 12.2 (30.4) | 10.1 (29.1) | 0.14 | 12.2 (30.5) | 10.1 (29.1) | 0.15 | 13.9 (32.2) | 8.6 (27.2) | <0.01 |
| **Water lead releases (lbs)** | 1.6 (7.2) | 1.6 (7.2) | 1.5 (7.3) | 0.72 | 1.7 (7.8) | 1.4 (6.7) | 0.30 | 1.5 (6.9) | 1.6 (7.6) | 0.84 | 1.8 (7.8) | 1.3 (6.7) | 0.15 |
| **Land Lead releases (lbs)** | 10.4 (232.5) | 4.9 (104.9) | 15.3 (305.7) | 0.35 | 1.5 (17.3) | 19.3 (328.4) | 0.12 | 19.3 (330.4) | 1.6 (17.6) | 0.12 | 4.5 (105.7) | 15.5 (302.5) | 0.31 |
| **Age at interview** | 76.1 (7.1) | 78.6 (7.6) | 73.7 (5.8) | <0.01 | 78.2 (7.6) | 73.9 (5.9) | <0.01 | 78.3 (7.6) | 73.9 (5.9) | <0.01 | 78.0 (7.6) | 74.4 (6.2) | <0.01 |
| **Sex** | | | | <0.01 | | | <0.01 | | | <0.01 | | | <0.01 |
| Male | 674 (41.1%) | 349 (44.6%) | 325 (38.0%) | | 383 (46.7%) | 291 (35.6%) | | 419 (51.9%) | 255 (30.7%) | | 251 (32.9%) | 423 (48.4%) | |
| Female | 964 (58.9%) | 434 (55.4%) | 530 (62.0%) | | 437 (53.3%) | 527 (64.4%) | | 389 (48.1%) | 575 (69.3%) | | 513 (67.1%) | 451 (51.6%) | |
| **Race/ethnicity** | | | | <0.01 | | | <0.01 | | | <0.01 | | | <0.01 |
| Asian | 410 (25.0%) | 211 (26.9%) | 199 (23.3%) | | 230 (28.0%) | 180 (22.0%) | | 169 (20.9%) | 241 (29.0%) | | 229 (30.0%) | 181 (20.7%) | |
| Black | 422 (25.8%) | 265 (33.8%) | 157 (18.4%) | | 254 (31.0%) | 168 (20.5%) | | 234 (29.0%) | 188 (22.7%) | | 299 (39.1%) | 123 (14.1%) | |
| LatinX | 322 (19.7%) | 172 (22.0%) | 150 (17.5%) | | 193 (23.5%) | 129 (15.8%) | | 185 (22.9%) | 137 (16.5%) | | 137 (17.9%) | 185 (21.2%) | |
| White | 484 (29.5%) | 135 (17.2%) | 349 (40.8%) | | 143 (17.4%) | 341 (41.7%) | | 220 (27.2%) | 264 (31.8%) | | 99 (13.0%) | 385 (44.1%) | |
| **Education** | | | | <0.01 | | | <0.01 | | | <0.01 | | | <0.01 |

| | | | | | | | | | | | | | |
|---|---|---|---|---|---|---|---|---|---|---|---|---|---|
| ≤ High School | 274 (16.7%) | 206 (26.3%) | 68 (8.0%) | | 206 (25.1%) | 68 (8.3%) | | 195 (24.1%) | 79 (9.5%) | | 190 (24.9%) | 84 (9.6%) | |
| > High School | 1364 (83.3%) | 577 (73.7%) | 787 (92.0%) | | 614 (74.9%) | 750 (91.7%) | | 613 (75.9%) | 751 (90.5%) | | 574 (75.1%) | 790 (90.4%) | |
| **Marital Status** | | | | <0.01 | | | <0.01 | | | <0.01 | | | <0.01 |
| Married/ living as married | 933 (57.0%) | 399 (51.0%) | 534 (62.5%) | | 434 (52.9%) | 499 (61.0%) | | 442 (54.7%) | 491 (59.2%) | | 349 (45.7%) | 584 (66.8%) | |
| Not married | 705 (43.0%) | 384 (49.0%) | 321 (37.5%) | | 386 (47.1%) | 319 (39.0%) | | 366 (45.3%) | 339 (40.8%) | | 415 (54.3%) | 290 (33.2%) | |
| **Average census tract income** | 121,338 (52,304) | 112,688 (4,9341) | 129,259 (53,696) | <0.01 | 114,432 (49,795) | 128,260 (53,852) | <0.01 | 117,017 (53,309) | 125,544 (50,987) | <0.01 | 112,006 (47,631) | 129,495 (54,810) | <0.01 |
| **Smoking status** | | | | 0.74 | | | 0.65 | | | 0.01 | | | 0.74 |
| Never | 912 (55.7%) | 433 (55.3%) | 479 (56.0%) | | 466 (56.8%) | 446 (54.5%) | | 424 (52.5%) | 488 (58.8%) | | 444 (58.1%) | 468 (53.5%) | |
| Past | 677 (41.3%) | 324 (41.4%) | 353 (41.3%) | | 330 (40.2%) | 347 (42.4%) | | 354 (43.8%) | 323 (38.9%) | | 298 (39.0%) | 379 (43.4%) | |
| Current | 49 (3.0%) | 26 (3.3%) | 23 (2.7%) | | 24 (2.9%) | 25 (3.1%) | | 30 (3.7%) | 19 (2.3%) | | 22 (2.9%) | 27 (3.1%) | |
| **Alcohol consumption** | | | | <0.01 | | | <0.01 | | | <0.01 | | | <0.01 |
| Never | 481 (29.4%) | 294 (37.5%) | 187 (21.9%) | | 291 (35.5%) | 190 (23.2%) | | 275 (34.0%) | 206 (24.8%) | | 295 (38.6%) | 186 (21.3%) | |
| Less than once a week | 572 (34.9%) | 266 (34.0%) | 306 (35.8%) | | 296 (36.1%) | 276 (33.7%) | | 247 (30.6%) | 325 (39.2%) | | 278 (36.4%) | 294 (33.6%) | |
| 1 - 6 days per week | 425 (25.9%) | 150 (19.2%) | 275 (32.2%) | | 164 (20.0%) | 261 (31.9%) | | 196 (24.3%) | 229 (27.6%) | | 138 (18.1%) | 287 (32.8%) | |

| | | | | | | | | |
|---|---|---|---|---|---|---|---|---|
| Every day | 160 (9.8%) | 73 (9.3%) | 87 (10.2%) | 69 (8.4%) | 91 (11.1%) | 90 (11.1%) | 70 (8.4%) | 53 (6.9%) | 107 (12.2%) |

Mean (SD) for continuous variables, N(%) categorical. [1]Fishers exact test, Satterthwaite t-test. [2]Cognitive measures are z-score standardized. [3]Total lead releases is a combination of air, land, water, and off-site lead releases reported by the facility to the Toxics Release Inventory. Off-site releases were not included in this analysis and therefore when combined, proportions of lead released through air, water, and land may not add to the total lead release number listed. Average census tract income rounded to the nearest whole number. Each cognitive outcome, dichotomized into “high” and “low” based on a cut-point of zero. lbs: pounds, km: kilometers.

**Supplemental Table 3.** Study of Healthy Aging in African Americans (STAR) analytic sample descriptive statistics by baseline cognitive domain test results.

| Characteristic | Overall | Cognitive measure | | | | | | | | | | | |
|---|---|---|---|---|---|---|---|---|---|---|---|---|---|
| | | Global cognition | | | Executive functioning | | | Episodic memory | | | Semantic memory | | |
| | N = 741 | Low N = 341 | High N = 400 | *p* value[1] | Low N = 354 | High N = 387 | *p* value[1] | Low N = 358 | High N = 383 | *p* value[1] | Low N = 350 | High N = 391 | *p* value[1] |
| **Average distance to facility (km)** | 3.6 (2.8) | 3.3 (1.9) | 3.8 (3.3) | 0.02 | 3.3 (1.9) | 3.8 (3.4) | <0.01 | 3.4 (1.9) | 3.7 (3.4) | 0.05 | 3.5 (3.3) | 3.7 (2.2) | 0.34 |
| **Lead facility within 1.5 km** | | | | 0.03 | | | 0.04 | | | 0.03 | | | 0.06 |
| No | 657 (88.7%) | 293 (85.9%) | 364 (91.0%) | | 305 (86.2%) | 352 (91.0%) | | 308 (86.0%) | 349 (91.1%) | | 302 (86.3%) | 355 (90.8%) | |
| Yes | 84 (11.3%) | 48 (14.1%) | 36 (9.0%) | | 49 (13.8%) | 35 (9.0%) | | 50 (14.0%) | 34 (8.9%) | | 48 (13.7%) | 36 (9.2%) | |
| **Lead facility within 3 km** | | | | 0.05 | | | <0.01 | | | 0.05 | | | 0.20 |
| No | 398 (53.7%) | 170 (49.9%) | 228 (57.0%) | | 170 (48.0%) | 228 (58.9%) | | 179 (50.0%) | 219 (57.2%) | | 179 (51.1%) | 219 (56.0%) | |
| Yes | 343 (46.3%) | 171 (50.1%) | 172 (43.0%) | | 184 (52.0%) | 159 (41.1%) | | 179 (50.0%) | 164 (42.8%) | | 171 (48.9%) | 172 (44.0%) | |
| **Lead facility within 5 km** | | | | 0.03 | | | 0.01 | | | 0.23 | | | 0.04 |
| No | 121 (16.3%) | 45 (13.2%) | 76 (19.0%) | | 45 (12.7%) | 76 (19.6%) | | 52 (14.5%) | 69 (18.0%) | | 47 (13.4%) | 74 (18.9%) | |
| Yes | 620 (83.7%) | 296 (86.8%) | 324 (81.0%) | | 309 (87.3%) | 311 (80.4%) | | 306 (85.5%) | 314 (82.0%) | | 303 (86.6%) | 317 (81.1%) | |
| **Total lead releases (lbs)[3]** | 4,971.3 (6,251.5) | 5,116.9 (6,306.9) | 4,847.1 (6,208.9) | 0.55 | 5,230.8 (6,300.3) | 4,733.9 (6,205.1) | 0.28 | 4,836.9 (6,245.1) | 5,096.9 (6,262.9) | 0.57 | 5,029.4 (6,267.2) | 4,919.2 (6,244.9) | 0.81 |
| **Air lead releases (lbs)** | 27.1 (36.9) | 28.8 (38.0) | 25.6 (36.0) | 0.24 | 29.3 (38.3) | 25.0 (35.7) | 0.11 | 26.9 (37.4) | 27.2 (36.6) | 0.92 | 27.9 (37.6) | 26.3 (36.5) | 0.54 |
| **Water lead releases (lbs)** | 0.8 (3.3) | 0.7 (2.7) | 0.8 (3.7) | 0.52 | 0.7 (2.4) | 0.9 (3.9) | 0.39 | 0.8 (3.3) | 0.8 (3.2) | 0.96 | 0.7 (2.7) | 0.8 (3.7) | 0.53 |

| | | | | | | | | | | | | | |
|---|---|---|---|---|---|---|---|---|---|---|---|---|---|
| **Land lead releases (lbs)** | 0.2 (4.6) | 0.4 (6.8) | 0.03 (0.4) | 0.35 | 0.0 (0.0) | 0.4 (6.4) | 0.28 | 0.0 (0.0) | 0.4 (6.5) | 0.28 | 0.4 (6.7) | 0.02 (0.4) | 0.33 |
| **Age at interview** | 68.8 (8.8) | 72.6 (9.4) | 65.5 (6.8) | <0.01 | 72.4 (9.1) | 65.4 (7.1) | <0.01 | 71.9 (9.3) | 65.8 (7.1) | <0.01 | 71.3 (9.6) | 66.5 (7.4) | <0.01 |
| **Sex** | | | | <0.01 | | | <0.01 | | | <0.01 | | | 0.02 |
| Male | 233 (31.4%) | 126 (37.0%) | 107 (26.8%) | | 134 (37.9%) | 99 (25.6%) | | 153 (42.7%) | 80 (20.9%) | | 95 (27.1%) | 138 (35.3%) | |
| Female | 508 (68.6%) | 215 (63.0%) | 293 (73.3%) | | 220 (62.1%) | 288 (74.4%) | | 205 (57.3%) | 303 (79.1%) | | 255 (72.9%) | 253 (64.7%) | |
| **Education** | | | | <0.01 | | | <0.01 | | | <0.01 | | | <0.01 |
| ≤ High School | 135 (18.2%) | 97 (28.4%) | 38 (9.5%) | | 98 (27.7%) | 37 (9.6%) | | 92 (25.7%) | 43 (11.2%) | | 98 (28.0%) | 37 (9.5%) | |
| > High School | 606 (81.8%) | 244 (71.6%) | 362 (90.5%) | | 256 (72.3%) | 350 (90.4%) | | 266 (74.3%) | 340 (88.8%) | | 252 (72.0%) | 354 (90.5%) | |
| **Marital Status** | | | | 0.20 | | | 0.33 | | | 0.65 | | | 0.71 |
| Married/ living as married | 328 (44.3%) | 142 (41.6%) | 186 (46.5%) | | 150 (42.4%) | 178 (46.0%) | | 155 (43.3%) | 173 (45.2%) | | 152 (43.4%) | 176 (45.0%) | |
| Not married | 413 (55.7%) | 199 (58.4%) | 214 (53.5%) | | 204 (57.6%) | 209 (54.0%) | | 203 (56.7%) | 210 (54.8%) | | 198 (56.6%) | 215 (55.0%) | |
| **Average census tract income** | 103,014 (48,533) | 100,406 (47,559) | 105,238 (49,299) | 0.17 | 97,922 (47,265) | 107,672 (49,267) | <0.01 | 100,971 (48,290) | 104,924 (48,745) | 0.26 | 99,127 (45,689) | 106,494 (50,751) | <0.01 |
| **Smoking status** | | | | 0.08 | | | 0.03 | | | <0.01 | | | 0.42 |
| Never | 391 (52.8%) | 168 (49.3%) | 223 (55.8%) | | 175 (49.4%) | 216 (55.8%) | | 171 (47.8%) | 220 (57.4%) | | 181 (51.7%) | 210 (53.7%) | |
| Past | 307 (41.4%) | 156 (45.7%) | 151 (37.8%) | | 163 (46.0%) | 144 (37.2%) | | 173 (48.3%) | 134 (35.0%) | | 152 (43.4%) | 155 (39.6%) | |
| Current | 43 (5.8%) | 17 (5.0%) | 26 (6.5%) | | 16 (4.5%) | 27 (7.0%) | | 14 (3.9%) | 29 (7.6%) | | 17 (4.9%) | 26 (6.6%) | |
| **Alcohol consumption** | | | | <0.01 | | | <0.01 | | | 0.01 | | | <0.01 |
| Never | 270 (36.4%) | 160 (46.9%) | 110 (27.5%) | | 158 (44.6%) | 112 (28.9%) | | 151 (42.2%) | 119 (31.1%) | | 160 (45.7%) | 110 (28.1%) | |
| Less than once a week | 237 (32.0.%) | 101 (29.6%) | 136 (34.0%) | | 105 (29.7%) | 132 (34.1%) | | 108 (30.2%) | 129 (33.7%) | | 102 (29.1%) | 135 (34.5%) | |

| | | | | | | | | | |
|---|---|---|---|---|---|---|---|---|---|
| 1 - 6 days per week | 208 (28.1%) | 73 (21.4%) | 135 (33.8%) | 83 (23.4%) | 125 (32.3%) | 89 (24.9%) | 119 (31.1%) | 81 (23.1%) | 127 (32.5%) |
| Every day | 26 (3.5%) | 7 (2.1%) | 19 (4.8%) | 8 (2.3%) | 18 (4.7%) | 10 (2.8%) | 16 (4.2%) | 7 (2.0%) | 19 (4.9%) |

Mean (SD) for continuous variables, N(%) categorical. [1]Fishers exact test, Satterthwaite t-test. [2]Cognitive measures are z-score standardized.[3]Total lead releases is a combination of air, land, water, and off-site lead releases reported by the facility to the Toxics Release Inventory. Off-site releases was not included in this analysis and therefore when combined, proportions of lead released through air, water, and land may not add to the total lead release number listed. Average census tract income rounded to the nearest whole number. Each cognitive outcome, dichotomized into “high” and “low” based on a cut-point of zero. lbs: pounds, km: kilometers.

**Supplemental Table 4.** Adjusted linear regression associations between residential distance to lead releasing facility and baseline cognition in the Kaiser Healthy Aging and Diverse Life Experiences Study (KHANDLE) analytic sample (n=1,638). Cognitive measures are z-score standardized.

| Distance to lead facility | Cognitive measure | | | | | | | | | | | |
|---|---|---|---|---|---|---|---|---|---|---|---|---|
| | Episodic memory | | | Semantic memory | | | Executive function | | | Global cognition | | |
| | β | 95% CI | *p* value | β | 95% CI | *p* value | β | 95% CI | *p* value | β | 95% CI | *p* value |
| **Minimally adjusted[1]** | | | | | | | | | | | | |
| Continuous (per 5 km) | -0.08 | (-0.05, -0.11) | <0.01 | -0.09 | (-0.06, -0.12) | <0.01 | -0.04 | (-0.01, -0.08) | <0.01 | -0.07 | (-0.05, -0.10) | <0.01 |
| Within 1.5 km | -0.21 | (-0.44, 0.02) | 0.07 | -0.34 | (-0.58, -0.10) | <0.01 | -0.44 | (-0.67, -0.20) | <0.01 | -0.33 | (-0.52, -0.14) | <0.01 |
| Within 3 km | -0.22 | (-0.33, -0.11) | <0.01 | -0.30 | (-0.41, -0.19) | <0.01 | -0.26 | (-0.37, -0.15) | <0.01 | -0.26 | (-0.35, -0.17) | <0.01 |
| Within 5 km | -0.25 | (-0.33, -0.16) | <0.01 | -0.27 | (-0.36, -0.18) | <0.01 | -0.16 | (-0.25, -0.07) | <0.01 | -0.23 | (-0.30, -0.15) | <0.01 |
| **Fully adjusted[2]** | | | | | | | | | | | | |
| Continuous (per 5 km) | -0.05 | (-0.02, -0.08) | <0.01 | -0.02 | (-0.05, 0.004) | 0.10 | 0.01 | (-0.02, 0.04) | 0.51 | -0.02 | (-0.04, 0.001) | 0.06 |
| Within 1.5 km | -0.09 | (-0.31, 0.13) | 0.43 | -0.16 | (-0.36, 0.03) | 0.11 | -0.24 | (-0.45, -0.04) | 0.02 | -0.17 | (-0.33, -0.01) | 0.04 |
| Within 3 km | -0.10 | (-0.21, 0.01) | 0.07 | -0.09 | (-0.19, 0.002) | 0.05 | -0.05 | (-0.15, 0.05) | 0.30 | -0.08 | (-0.16, -0.004) | 0.04 |
| Within 5 km | -0.15 | (-0.24, -0.06) | <0.01 | -0.07 | (-0.15, 0.01) | 0.07 | 0.002 | (-0.08, 0.08) | 0.99 | -0.07 | (-0.14, -0.01) | 0.02 |

[1]Minimally adjusted for baseline age and sex. [2] Fully adjusted for age, sex, education, income, marital status, race/ethnicity, smoking status, and alcohol consumption. Continuous distance interpreted per every 5 km closer a residence is to a lead facility. β: beta value for linear regression estimate, km: kilometers.

**Supplemental Table 5.** Adjusted linear regression associations between residential distance to lead releasing facility and baseline cognition in the Study of Healthy Aging in African Americans (STAR) analytic sample (n=741). Cognitive measures are z-score standardized.

| Distance to lead facility | Cognitive measure | | | | | | | | | | | |
|---|---|---|---|---|---|---|---|---|---|---|---|---|
| | Episodic memory | | | Semantic memory | | | Executive function | | | Global cognition | | |
| | β | 95% CI | P value | β | 95% CI | P value | β | 95% CI | P value | β | 95% CI | P value |
| **Minimally adjusted[1]** | | | | | | | | | | | | |
| Continuous (per 5 km) | -0.08 | (-0.20, 0.03) | 0.15 | -0.12 | (-0.004, -0.24) | 0.04 | -0.10 | (-0.21, 0.01) | 0.07 | -0.10 | (-0.01, -0.19) | 0.03 |
| Within 1.5 km | -0.20 | (-0.40, -0.005) | 0.04 | -0.17 | (-0.38, 0.04) | 0.11 | -0.16 | (-0.35, 0.04) | 0.12 | -0.18 | (-0.33, -0.02) | 0.03 |
| Within 3 km | -0.11 | (-0.23, 0.02) | 0.09 | -0.14 | (-0.27, -0.01) | 0.04 | -0.17 | (-0.30, -0.05) | <0.01 | -0.14 | (-0.24, -0.04) | <0.01 |
| Within 5 km | -0.09 | (-0.26, 0.08) | 0.32 | -0.28 | (-0.46, -0.10) | <0.01 | -0.10 | (-0.27, 0.07) | 0.23 | -0.16 | (-0.29, -0.02) | 0.02 |
| **Fully adjusted[2]** | | | | | | | | | | | | |
| Continuous (per 5 km) | -0.03 | (-0.15, 0.08) | 0.59 | -0.07 | (-0.19, 0.05) | 0.25 | -0.02 | (-0.13, 0.09) | 0.67 | -0.04 | (-0.13, 0.05) | 0.35 |
| Within 1.5 km | -0.13 | (-0.33, 0.07) | 0.19 | -0.08 | (-0.28, 0.13) | 0.46 | -0.05 | (-0.23, 0.14) | 0.64 | -0.08 | (-0.23, 0.07) | 0.28 |
| Within 3 km | -0.06 | (-0.19, 0.07) | 0.37 | -0.10 | (-0.24, 0.04) | 0.16 | -0.09 | (-0.22, 0.04) | 0.19 | -0.08 | (-0.18, 0.02) | 0.12 |
| Within 5 km | 0.02 | (-0.16, 0.21) | 0.79 | -0.20 | (-0.39, -0.02) | 0.03 | 0.08 | (-0.10, 0.26) | 0.37 | -0.03 | (-0.17, 0.11) | 0.64 |

[1]Minimally adjusted for baseline age and sex. [2]Fully adjusted for age, sex, education, income, marital status, race/ethnicity, smoking status, and alcohol consumption. Continuous distance interpreted per every 5 km closer a residence is to a lead facility. β: beta value for linear regression estimate, km: kilometers.

**Supplemental Table 6.** Table of mean difference and 95% confidence interval for meta-analysis across the KHANDLE and STAR analytic sample cohorts.

| Distance to lead facility | Cognitive measure | | | | | | | | | | | |
|---|---|---|---|---|---|---|---|---|---|---|---|---|
| | Episodic memory | | | Semantic memory | | | Executive function | | | Global cognition | | |
| | MD | 95% CI | P value | MD | 95% CI | P value | MD | 95% CI | P value | MD | 95% CI | P value |
| **Minimally adjusted[1]** | | | | | | | | | | | | |
| Continuous (per 5 km) | -0.08 | (-0.11, -0.05) | <0.01 | -0.09 | (-0.12, -0.06) | <0.01 | -0.05 | (-0.08, -0.02) | <0.01 | -0.07 | (-0.10, -0.05) | <0.01 |
| Within 1.5 km | -0.21 | (-0.36, -0.06) | <0.01 | -0.24 | (-0.41, -0.08) | <0.01 | -0.29 | (-0.56,-0.01) | 0.04 | -0.24 | (-0.39,-0.09) | <0.01 |
| Within 3 km | -0.17 | (-0.28, -0.06) | <0.01 | -0.23 | (-0.38, -0.07) | <0.01 | -0.22 | (-0.30, -0.14) | <0.01 | -0.20 | (-0.32, -0.09) | <0.01 |
| Within 5 km | -0.18 | (-0.34, -0.03) | 0.02 | -0.27 | (-0.35, -0.19) | <0.01 | -0.15 | (-0.23, -0.07) | <0.01 | -0.21 | (-0.27, -0.15) | <0.01 |
| **Fully adjusted[2]** | | | | | | | | | | | | |
| Continuous (per 5 km) | -0.05 | (-0.08, -0.02) | <0.01 | -0.03 | (-0.05, -0.001) | 0.06 | 0.008 | (-0.02, 0.04) | 0.60 | -0.02 | (-0.04, -0.0008) | 0.04 |
| Within 1.5 km | -0.11 | (-0.26, 0.03) | 0.13 | -0.12 | (-0.26, 0.02) | 0.10 | -0.14 | (-0.33, 0.05) | 0.16 | -0.12 | (-0.23,-0.01) | 0.03 |
| Within 3 km | -0.08 | (-0.17, 0.0001) | 0.05 | -0.10 | (-0.18, -0.02) | 0.02 | -0.06 | (-0.15, 0.01) | 0.10 | -0.08 | (-0.14, -0.02) | <0.01 |
| Within 5 km | -0.08 | (-0.25, 0.08) | 0.33 | -0.11 | (-0.23, 0.01) | 0.06 | 0.02 | (-0.06,0.09) | 0.68 | -0.07 | (-0.12, -0.009) | 0.02 |

[1]Minimally adjusted for age and sex. [2]Fully adjusted for age, sex, education, income, marital status, race/ethnicity, smoking status, alcohol consumption. KHANDLE: Kaiser Healthy Aging and Diverse Life Experiences Study; STAR: Study of Healthy Aging in African Americans, MD: mean difference.

**Supplemental Table 7.** Descriptive statistics for the complete case sensitivity analysis included and excluded participants in the KHANDLE and STAR cohorts.

| | Cohort | | | | | | | |
|---|---|---|---|---|---|---|---|---|
| | KHANDLE | | | | STAR | | | |
| **Characteristic** | **Overall,** N = 1,663 | **Excluded,** N = 73 | **Included,** N = 1,590 | ***p* value**[1] | **Overall,** N = 746 | **Excluded,** N = 25 | **Included,** N = 721 | ***p* value**[1] |
| **Global cognition**[2] | 0.01 (0.81) | -0.22 (0.91) | 0.02 (0.81) | 0.07 | 0.01 (0.81) | -0.51 (0.90) | 0.03 (0.80) | 0.02 |
| (missing) | 1 | 1 | - | | - | - | - | |
| **Executive function**[2] | 0.01 (1.00) | -0.38 (0.99) | 0.02 (0.99) | <0.01 | 0.01 (0.99) | -0.73 (.99) | 0.03 (0.99) | <0.01 |
| (missing) | 10 | 10 | - | | - | - | - | |
| **Episodic memory**[2] | 0.01 (1.00) | -0.08 (1.20) | 0.01 (0.99) | 0.60 | 0.02 (0.99) | -0.37 (1.00) | 0.03 (0.99) | 0.08 |
| (missing) | 4 | 4 | - | | 2 | 2 | - | |
| **Semantic memory**[2] | 0.01 (1.00) | -0.27 (1.10) | 0.02 (0.99) | 0.06 | 0.00 (0.99) | -0.50 (1.10) | 0.02 (0.98) | 0.03 |
| (missing) | 7 | 7 | - | | 3 | 3 | - | |
| **Average distance to facility (km)** | 8.2 (6.9) | 7.5 (6.4) | 8.2 (6.9) | 0.36 | 3.6 (2.8) | 3.4 (1.1) | 3.6 (2.8) | 0.40 |
| **Lead releasing facility within 1.5km** | | | | 0.19 | | | | 0.34 |
| No | 1602 (96.3%) | 68 (93.2%) | 1,534 (96.5%) | | 661 (88.6%) | 22 (95.7%) | 637 (88.3%) | |
| Yes | 61 (3.7%) | 5 (6.8%) | 56 (3.5%) | | 85 (11.4%) | 1 (4.3%) | 84 (11.7%) | |
| **Lead releasing facility within 3km** | | | | 0.46 | | | | 0.16 |
| No | 1331 (80.0%) | 56 (76.7%) | 1,275 (80.2%) | | 401 (53.8%) | 15 (65.2%) | 384 (53.3%) | |
| Yes | 332 (20.0%) | 17 (23.3%) | 315 (19.8%) | | 345 (46.2%) | 8 (34.8%) | 337 (46.7%) | |
| **Lead releasing facility within 5km** | | | | 0.23 | | | | 0.16 |
| No | 907 (54.5%) | 37 (50.7%) | 870 (54.7%) | | 122 (16.4%) | - | 66 (9.2%) | |

| | | | | | | | | |
|---|---|---|---|---|---|---|---|---|
| Yes | 756 (45.5%) | 45 (61.6%) | 862 (54.2%) | | 624 (83.6%) | 23 (100.0%) | 655 (90.8%) | |
| **Total lead releases (lbs)[3]** | 2,307.5 (5,331.9) | 2,379.1 (4,899.1) | 2,304.2 (5,352.3) | 0.90 | 4,956.1 (6,245.5) | 3,516.3 (5,831.1) | 5,006.1 (6,257.2) | 0.20 |
| **Air lead releases (lbs)** | 11.3 (30.1) | 15.5 (35.2) | 11.1 (29.9) | 0.30 | 27.0 (37.0) | 19.8 (34.2) | 27.2 (37.1) | 0.30 |
| **Water lead releases (lbs)** | 1.5 (7.2) | 2.2 (9.9) | 1.5 (7.0) | 0.56 | 0.8 (3.3) | 0.2 (0.7) | 0.8 (3.3) | <0.01 |
| **Land lead releases (lbs)** | 10.2 (230.7) | 0.0 | 10.7 (236.0) | 0.07 | 0.2 (4.6) | 0.0 (0.0) | 0.2 (4.7) | 0.30 |
| **Age at interview** | 76.1 (7.2) | 78.7 (8.8) | 76 (7.1) | 0.01 | 68.8 (8.8) | 69.7 (7.8) | 68.7 (8.8) | 0.50 |
| **Sex** | | | | 0.90 | | | | 0.20 |
| Male | 684 (41.1%) | 29 (39.7%) | 655 (41.2%) | | 235 (31.5%) | 11 (47.8%) | 224 (31.1%) | |
| Female | 979 (58.9%) | 44 (60.3%) | 935 (58.8%) | | 511 (68.5%) | 12 (52.2%) | 497 (68.9%) | |
| **Race/ethnicity** | | | | <0.01 | | | | <0.01 |
| Asian | 412 (24.8%) | 15 (20.5%) | 397 (25.0%) | | - | - | - | |
| Black | 431 (25.9%) | 23 (31.5%) | 408 (25.7%) | | 736 (98.7%) | 19 (82.6%) | 715 (99.2%) | |
| LatinX | 327 (19.7%) | 13 (17.8%) | 314 (19.7%) | | 6 (0.8%) | - | 6 (0.8%) | |
| Native American | 3 (0.2%) | 3 (4.1%) | - | | 4 (0.5%) | 4 (17.4%) | - | |
| White | 490 (29.5%) | 19 (26.0%) | 471 (29.6%) | | - | - | - | |
| (missing) | 1 | 1 | - | | | | | |
| **Education** | | | | <0.01 | | | | <0.01 |
| ≤ High School | 281 (16.9%) | 18 (24.7%) | 262 (16.5%) | | 136 (18.2%) | 7 (30.4%) | 128 (17.8%) | |
| > High School | 1382 (83.1%) | 54 (74.0%) | 1,328 (83.5%) | | 610 (81.8%) | 13 (56.5%) | 593 (82.2%) | |
| (missing) | 1 | 1 | - | | 3 | 3 | - | |
| **Marital Status** | | | | <0.01 | | | | <0.01 |
| Married/ living as married | 944 (56.8%) | 25 (34.2%) | 908 (57.1%) | | 329 (44.1%) | 1 (4.3%) | 320 (44.4%) | |
| Not Married | 714 (42.9%) | 16 (21.9%) | 682 (42.9%) | | 416 (55.8%) | 5 (21.7%) | 401 (55.6%) | |

| | | | | | | | | |
|---|---|---|---|---|---|---|---|---|
| (missing) | 26 | 26 | - | | 17 | 17 | - | |
| **Average census tract income** | 1,213,78 (52,401) | 118,853 (52,807) | 121,502 (52,344) | 0.68 | 103,178 (48,673) | 100,955 (45,476) | 103,255 (48,808) | 0.80 |
| (missing) | 3 | 3 | - | | - | - | - | |
| **Smoking status** | | | | <0.01 | | | | 0.74 |
| Never | 920 (55.3%) | 32 (43.8%) | 887 (55.8%) | | 396 (53.1%) | 13 (56.5%) | 382 (53.0%) | |
| Former | 691 (41.6%) | 31 (42.5%) | 656 (41.3%) | | 307 (41.2%) | 8 (34.8%) | 298 (41.3%) | |
| Current | 52 (3.1%) | 4 (5.5%) | 47 (3.0%) | | 43 (5.8%) | 2 (8.7%) | 41 (5.7%) | |
| (missing) | 6 | 6 | - | | - | - | - | |
| **Alcohol consumption** | | | | <0.01 | | | | <0.01 |
| Never | 492 (29.6%) | 24 (32.9%) | 466 (29.3%) | | 274 (36.7%) | 15 (65.2%) | 259 (35.9%) | |
| Less than once a week | 580 (34.9%) | 21 (28.8%) | 551 (34.7%) | | 237 (31.8%) | 3 (13.0%) | 232 (32.2%) | |
| 1-6 days per week | 430 (25.9%) | 14 (19.2%) | 414 (26.0%) | | 208 (27.9%) | 4 (17.4%) | 204 (28.3%) | |
| Every day | 161 (9.7%) | 1 (1.4%) | 159 (10.0%) | | 27 (3.6%) | 1 (4.3%) | 26 (3.6%) | |
| (missing) | 12 | 12 | - | | - | - | - | |

Mean (SD) for continuous variables, N(%) categorical. [1]Fishers exact test, Satterthwaite t-test. [2]Cognitive measures are z-score standardized. [3]Total lead releases is a combination of air, land, water, and off-site lead releases reported by the facility to the Toxics Release Inventory. Off-site releases was not included in this analysis and therefore when combined, proportions of lead released through air, water, and land may not add to the total lead release number listed. Average census tract income rounded to the nearest whole number, KHANDLE: Kaiser Healthy Aging and Diverse Life Experiences Study, STAR: Study of Healthy Aging in African Americans, lbs: pounds, km: kilometers.

**Supplemental Table 8**. Sensitivity results for complete case analysis adjusted linear regression associations between residential distance to lead releasing facility and baseline cognition in KHANDLE analytic sample (n=1,590). Cognitive measures are z-score standardized.

| Distance to lead facility | Cognitive measure | | | | | | | | | | | |
|---|---|---|---|---|---|---|---|---|---|---|---|---|
| | Episodic memory | | | Semantic memory | | | Executive function | | | Global cognition | | |
| | β | 95% CI | P value | β | 95% CI | P value | β | 95% CI | P value | β | 95% CI | P value |
| **Minimally adjusted**[1] | | | | | | | | | | | | |
| Continuous (per 5 km) | -0.07 | (-0.04, -0.10) | <0.01 | -0.09 | (-0.05, -0.12) | <0.01 | -0.04 | (-0.01, -0.07) | 0.01 | -0.07 | (-0.04, -0.09) | <0.01 |
| Within 1.5 km | -0.16 | (-0.40, 0.08) | 0.19 | -0.31 | (-0.56, -0.07) | 0.01 | -0.38 | (-0.63, -0.14) | <0.01 | -0.28 | (-0.48, -0.09) | <0.01 |
| Within 3 km | -0.20 | (-0.31, -0.10) | <0.01 | -0.28 | (-0.40, -0.17) | <0.01 | -0.25 | (-0.36, -0.13) | <0.01 | -0.25 | (-0.34, -0.16) | <0.01 |
| Within 5 km | -0.22 | (-0.31, -0.14) | <0.01 | -0.26 | (-0.35, -0.17) | <0.01 | -0.16 | (-0.25, -0.07) | <0.01 | -0.21 | (-0.29, -0.14) | <0.01 |
| **Fully adjusted**[2] | | | | | | | | | | | | |
| Continuous (per 5 km) | -0.04 | (-0.01, -0.08) | 0.01 | -0.02 | (-0.04, 0.01) | 0.17 | 0.01 | (-0.02, 0.04) | 0.45 | -0.02 | (-0.04, 0.01) | 0.13 |
| Within 1.5 km | -0.05 | (-0.28, 0.18) | 0.69 | -0.17 | (-0.37, 0.04) | 0.11 | -0.21 | (-0.43, 0.002) | 0.05 | -0.14 | (-0.31, 0.02) | 0.09 |
| Within 3 km | -0.09 | (-0.2, 0.02) | 0.11 | -0.08 | (-0.18, 0.02) | 0.11 | -0.05 | (-0.15, 0.06) | 0.38 | -0.07 | (-0.15, 0.01) | 0.08 |
| Within 5 km | -0.13 | (-0.22, -0.05) | <0.01 | -0.07 | (-0.15, 0.01) | 0.09 | <0.01 | (-0.08, 0.09) | 0.94 | -0.07 | (-0.13, -0.002) | 0.04 |

[1]Minimally adjusted for age and sex. [2]Fully adjusted for age, sex, education, income, marital status, race/ethnicity, smoking status, alcohol consumption. Complete case: non-missing values for all observations required for study inclusion. Continuous distance interpreted per every 5 km closer a residence is to a lead facility. β: beta value for linear regression estimate, km: kilometers, KHANDLE: Kaiser Healthy Aging and Diverse Life Experiences Study.

**Supplemental Table 9**. Sensitivity results for complete case analysis adjusted linear regression associations between residential distance to lead releasing facility and baseline cognition in STAR analytic sample (n=721). Cognitive measures are z-score standardized.

| Distance to lead facility | Cognitive measure | | | | | | | | | | | |
|---|---|---|---|---|---|---|---|---|---|---|---|---|
| | Episodic memory | | | Semantic memory | | | Executive function | | | Global cognition | | |
| | **β** | **95% CI** | **P value** | **β** | **95% CI** | **P value** | **β** | **95% CI** | **P value** | **β** | **95% CI** | **P value** |
| **Minimally adjusted**[1] | | | | | | | | | | | | |
| Continuous (per 5 km) | -0.08 | (-0.19, 0.03) | 0.16 | -0.13 | (-0.01, -0.25) | 0.03 | -0.10 | (-0.21, 0.01) | 0.07 | -0.10 | (-0.02, -0.19) | 0.02 |
| Within 1.5 km | -0.21 | (-0.41, -0.01) | 0.04 | -0.19 | (-0.39, 0.02) | 0.07 | -0.17 | (-0.37, 0.02) | 0.08 | -0.19 | (-0.35, -0.04) | 0.02 |
| Within 3 km | -0.12 | (-0.25, 0.01) | 0.06 | -0.18 | (-0.31, -0.05) | 0.01 | -0.19 | (-0.32, -0.07) | <0.01 | -0.16 | (-0.26, -0.07) | <0.01 |
| Within 5 km | -0.08 | (-0.25, 0.09) | 0.38 | -0.26 | (-0.44, -0.09) | <0.01 | -0.08 | (-0.25, 0.08) | 0.33 | -0.14 | (-0.28, -0.01) | 0.04 |
| **Fully adjusted**[2] | | | | | | | | | | | | |
| Continuous (per 5 km) | -0.03 | (-0.14, 0.08) | 0.6 | -0.08 | (-0.20, 0.04) | 0.19 | 0.02 | (-0.13, 0.09) | 0.66 | -0.04 | (-0.13, 0.04) | 0.32 |
| Within 1.5 km | -0.14 | (-0.33, 0.06) | 0.17 | -0.09 | (-0.30, 0.11) | 0.36 | -0.06 | (-0.25, 0.13) | 0.52 | -0.10 | (-0.25, 0.05) | 0.20 |
| Within 3 km | -0.07 | (-0.20, 0.07) | 0.34 | -0.14 | (-0.28, 0.003) | 0.05 | -0.10 | (-0.23, 0.03) | 0.13 | -0.10 | (-0.21, 0.002) | 0.05 |
| Within 5 km | 0.03 | (-0.15, 0.22) | 0.72 | -0.19 | (-0.38, -0.004) | 0.05 | 0.10 | (-0.07, 0.28) | 0.26 | -0.02 | (-0.16, 0.12) | 0.79 |

[1]Minimally adjusted for age and sex. [2]Fully adjusted for age, sex, education, income, marital status, race/ethnicity, smoking status, alcohol consumption. Complete case: non-missing values for all observations required for study inclusion. Continuous distance interpreted per every 5 km closer a residence is to a lead facility. β: beta value for linear regression estimate, km: kilometers, STAR: Study of Healthy Aging in African Americans.

**Supplemental Table 10:** Sensitivity results for age stratified analysis adjusted linear regression associations between residential distance to lead releasing facility and baseline cognition in KHANDLE analytic sample (n=1,638). Cognitive measures are z-score standardized.

| Distance to lead facility | Age | Cognitive measure | | | | | | | | | | | |
|---|---|---|---|---|---|---|---|---|---|---|---|---|---|
| | | Episodic memory | | | Semantic memory | | | Executive function | | | Global cognition | | |
| | | β | 95% CI | P value | β | 95% CI | P value | β | 95% CI | P value | β | 95% CI | P value |
| **Minimally adjusted**[1] | | | | | | | | | | | | | |
| Continuous (per 5 km) | < 70 | -0.09 | (-0.01, -0.16) | 0.03 | -0.10 | (-0.02, -0.17) | 0.01 | -0.07 | (-0.14, 0.01) | 0.08 | -0.08 | (-0.02, -0.15) | 0.01 |
| | 70-74 | -0.10 | (-0.05, -0.16) | <0.01 | -0.04 | (-0.02, 0.11) | 0.17 | -0.04 | (-0.10, 0.02) | 0.24 | 0.06 | (0.01, 0.11) | 0.01 |
| | 75-79 | -0.05 | (-0.11, 0.01) | 0.12 | -0.12 | (0.05, 0.19) | <0.01 | -0.05 | (-0.12, 0.02) | 0.15 | 0.07 | (0.02, 0.13) | 0.01 |
| | ≥ 80 | -0.08 | (-0.02, -0.13) | 0.01 | -0.10 | (0.04, 0.16) | <0.01 | -0.03 | (-0.09, 0.03) | 0.29 | 0.07 | (0.02, 0.12) | <0.01 |
| Within 1.5 km | < 70 | -0.05 | (-0.53, 0.43) | 0.84 | -0.05 | (-0.51, 0.41) | 0.83 | -0.17 | (-0.63, 0.30) | 0.48 | -0.09 | (-0.47, 0.29) | 0.64 |
| | 70-74 | 0.07 | (-0.36, 0.49) | 0.75 | -0.62 | (-1.08, -0.17) | 0.01 | -0.32 | (-0.77, 0.14) | 0.17 | -0.29 | (-0.65, 0.07) | 0.11 |
| | 75-79 | -0.50 | (-0.91, -0.09) | 0.02 | -0.50 | (-0.93, -0.06) | 0.02 | -0.79 | (-1.22, -0.36) | <0.01 | -0.60 | (-0.93, -0.26) | <0.01 |
| | ≥ 80 | -0.47 | (-1.07, 0.12) | 0.12 | -0.07 | (-0.70, 0.56) | 0.83 | -0.57 | (-1.18, 0.04) | 0.07 | -0.37 | (-0.87, 0.13) | 0.14 |
| Within 3 km | < 70 | -0.07 | (-0.30, 0.15) | 0.53 | -0.07 | (-0.29, 0.14) | 0.51 | -0.07 | (-0.29, 0.15) | 0.51 | -0.07 | (-0.25, 0.10) | 0.42 |
| | 70-74 | -0.32 | (-0.52, -0.12) | <0.01 | -0.29 | (-0.51, -0.08) | 0.01 | -0.23 | (-0.44, -0.02) | 0.03 | -0.28 | (-0.45, -0.12) | <0.01 |
| | 75-79 | -0.23 | (-0.45, 0.004) | 0.05 | -0.38 | (-0.62, -0.14) | <0.01 | -0.31 | (-0.55, -0.07) | 0.01 | -0.31 | (-0.49, -0.12) | <0.01 |
| | ≥ 80 | -0.33 | (-0.55, -0.11) | <0.01 | -0.54 | (-0.77, -0.31) | <0.01 | -0.52 | (-0.74, -0.29) | <0.01 | -0.46 | (-0.64, -0.28) | <0.01 |
| Within 5 km | < 70 | -0.12 | (-0.31, 0.06) | 0.19 | -0.28 | (-0.45, -0.10) | <0.01 | -0.11 | (-0.29, 0.08) | 0.26 | -0.17 | (-0.31, -0.02) | 0.02 |
| | 70-74 | -0.32 | (-0.47, -0.16) | <0.01 | -0.04 | (-0.22, 0.13) | 0.61 | -0.11 | (-0.28, 0.06) | 0.19 | -0.16 | (-0.29, -0.02) | 0.02 |
| | 75-79 | -0.17 | (-0.36, 0.03) | 0.09 | -0.48 | (-0.68, -0.28) | <0.01 | -0.29 | (-0.49, -0.08) | 0.01 | -0.31 | (-0.47, -0.15) | <0.01 |
| | ≥ 80 | -0.35 | (-0.51, -0.18) | <0.01 | -0.35 | (-0.53, -0.17) | <0.01 | -0.20 | (-0.37, -0.03) | 0.02 | -0.30 | (-0.44, -0.16) | <0.01 |
| **Fully adjusted**[2] | | | | | | | | | | | | | |
| Continuous (per 5 km) | < 70 | -0.04 | (-0.12, 0.03) | 0.25 | -0.03 | (-0.10, 0.03) | 0.28 | -0.03 | (-0.09, 0.04) | 0.44 | -0.04 | (-0.09, 0.02) | 0.19 |

| | | | | | | | | | | | | | |
|---|---|---|---|---|---|---|---|---|---|---|---|---|---|
| | 70-74 | -0.07 | (-0.02, -0.13) | 0.01 | 0.01 | (-0.05, 0.06) | 0.75 | 0.01 | (-0.04, 0.07) | 0.65 | -0.02 | (-0.06, 0.03) | 0.44 |
| | 75-79 | -0.03 | (-0.09, 0.03) | 0.36 | -0.04 | (-0.09, 0.02) | 0.18 | 0.02 | (-0.04, 0.08) | 0.53 | -0.02 | (-0.06, 0.03) | 0.47 |
| | ≥ 80 | -0.05 | (-0.11, 0.01) | 0.09 | -0.03 | (-0.08, 0.02) | 0.25 | 0.02 | (-0.03, 0.08) | 0.39 | -0.02 | (-0.06, 0.02) | 0.39 |
| Within 1.5 km | < 70 | 0.01 | (-0.44, 0.46) | 0.95 | 0.19 | (-0.18, 0.55) | 0.31 | 0.02 | (-0.38, 0.42) | 0.93 | 0.07 | (-0.24, 0.38) | 0.64 |
| | 70-74 | 0.29 | (-0.11, 0.69) | 0.16 | -0.37 | (-0.76, 0.01) | 0.06 | -0.05 | (-0.44, 0.35) | 0.82 | -0.04 | (-0.35, 0.26) | 0.78 |
| | 75-79 | -0.38 | (-0.78, 0.02) | 0.06 | -0.26 | (-0.61, 0.09) | 0.15 | -0.50 | (-0.86, -0.14) | 0.01 | -0.38 | (-0.66, -0.10) | 0.01 |
| | ≥ 80 | -0.31 | (-0.89, 0.28) | 0.30 | -0.003 | (-0.53, 0.52) | 0.99 | -0.47 | (-1.01, 0.08) | 0.10 | -0.26 | (-0.69, 0.18) | 0.25 |
| Within 3 km | < 70 | -0.03 | (-0.25, 0.19) | 0.81 | 0.06 | (-0.12, 0.23) | 0.54 | 0.05 | (-0.15, 0.24) | 0.64 | 0.03 | (-0.13, 0.18) | 0.74 |
| | 70-74 | -0.18 | (-0.38, 0.01) | 0.07 | -0.05 | (-0.24, 0.14) | 0.60 | 0.01 | (-0.18, 0.21) | 0.89 | -0.07 | (-0.22, 0.08) | 0.34 |
| | 75-79 | -0.05 | (-0.29, 0.19) | 0.68 | -0.04 | (-0.25, 0.17) | 0.70 | 0.02 | (-0.19, 0.23) | 0.86 | -0.02 | (-0.19, 0.14) | 0.78 |
| | ≥ 80 | -0.18 | (-0.40, 0.05) | 0.12 | -0.33 | (-0.53, -0.12) | <0.01 | -0.32 | (-0.53, -0.10) | <0.01 | -0.27 | (-0.44, -0.11) | <0.01 |
| Within 5 km | < 70 | -0.06 | (-0.24, 0.12) | 0.49 | -0.16 | (-0.31, -0.02) | 0.03 | -0.04 | (-0.20, 0.12) | 0.64 | -0.09 | (-0.21, 0.03) | 0.16 |
| | 70-74 | -0.23 | (-0.39, -0.08) | <0.01 | 0.04 | (-0.11, 0.19) | 0.56 | -0.03 | (-0.19, 0.12) | 0.66 | -0.07 | (-0.19, 0.04) | 0.22 |
| | 75-79 | -0.08 | (-0.28, 0.13) | 0.46 | -0.13 | (-0.31, 0.05) | 0.16 | 0.02 | (-0.16, 0.20) | 0.84 | -0.06 | (-0.21, 0.08) | 0.40 |
| | ≥ 80 | -0.22 | (-0.40, -0.04) | 0.02 | -0.09 | (-0.25, 0.07) | 0.29 | 0.04 | (-0.13, 0.21) | 0.67 | -0.09 | (-0.22, 0.04) | 0.19 |

[1]Minimally adjusted for baseline age and sex. [2]Fully adjusted for age, sex, education, income, marital status, race/ethnicity, smoking status, alcohol consumption. Continuous distance interpreted per every 5 km closer a residence is to a lead facility. β: beta value for linear regression estimate, km: kilometers, KHANDLE: Kaiser Healthy Aging and Diverse Life Experiences Study

**Supplemental Table 11:** Sensitivity results for age stratified adjusted linear regression associations between residential distance to lead releasing facility and baseline cognition in STAR analytic sample (n=741). Cognitive measures are z-score standardized.

| Distance to lead facility | Age | Cognitive measure | | | | | | | | | | | |
|---|---|---|---|---|---|---|---|---|---|---|---|---|---|
| | | Episodic memory | | | Semantic memory | | | Executive function | | | Global cognition | | |
| | | β | 95% CI | P value | β | 95% CI | P value | β | 95% CI | P value | β | 95% CI | P value |
| **Minimally adjusted** [1] | | | | | | | | | | | | | |
| Continuous (per 5 km) | < 70 | -0.04 | (-0.16, 0.08) | 0.54 | -0.07 | (-0.19, 0.05) | 0.23 | -0.05 | (-0.17, 0.07) | 0.38 | -0.06 | (-0.15, 0.04) | 0.25 |
| | 70-74 | 0.003 | (-0.40, 0.41) | 0.99 | -0.13 | (-0.53, 0.26) | 0.51 | -0.22 | (-0.60, 0.16) | 0.26 | -0.12 | (-0.42, 0.19) | 0.46 |
| | 75-79 | -0.91 | (-0.42, -1.40) | <0.01 | -0.43 | (-1.10, 0.22) | 0.20 | -0.46 | (-0.98, 0.05) | 0.08 | -0.60 | (-0.17, -1.03) | <0.01 |
| | ≥ 80 | -0.32 | (-0.88, 0.25) | 0.27 | -0.83 | (-0.23, -1.40) | <0.01 | -0.78 | (-0.19, -1.37) | 0.01 | -0.64 | (-0.15, -1.10) | 0.01 |
| Within 1.5 km | < 70 | -0.07 | (-0.33, 0.20) | 0.62 | -0.09 | (-0.35, 0.18) | 0.52 | 0.04 | (-0.23, 0.30) | 0.79 | -0.04 | (-0.24, 0.16) | 0.70 |
| | 70-74 | -0.43 | (-0.88, 0.02) | 0.06 | 0.26 | (-0.18, 0.71) | 0.25 | -0.46 | (-0.88, -0.03) | 0.04 | -0.21 | (-0.56, 0.14) | 0.23 |
| | 75-79 | -0.25 | (-0.86, 0.36) | 0.41 | -0.45 | (-1.21, 0.30) | 0.24 | -0.20 | (-0.81, 0.40) | 0.51 | -0.30 | (-0.82, 0.21) | 0.25 |
| | ≥ 80 | -0.36 | (-0.88, 0.16) | 0.17 | -0.92 | (-1.47, -0.37) | <0.01 | -0.56 | (-1.12, 0.0007) | 0.05 | -0.61 | (-1.07, -0.16) | 0.01 |
| Within 3 km | < 70 | -0.09 | (-0.25, 0.07) | 0.29 | -0.17 | (-0.32, -0.01) | 0.04 | -0.08 | (-0.24, 0.08) | 0.34 | -0.11 | (-0.23, 0.01) | 0.08 |
| | 70-74 | -0.11 | (-0.44, 0.23) | 0.53 | 0.02 | (-0.30, 0.35) | 0.89 | -0.34 | (-0.65, -0.03) | 0.03 | -0.14 | (-0.39, 0.11) | 0.27 |
| | 75-79 | -0.31 | (-0.69, 0.07) | 0.11 | -0.01 | (-0.49, 0.47) | 0.96 | -0.33 | (-0.70, 0.05) | 0.09 | -0.22 | (-0.54, 0.11) | 0.19 |
| | ≥ 80 | -0.23 | (-0.57, 0.11) | 0.19 | -0.56 | (-0.93, -0.20) | <0.01 | -0.60 | (-0.96, -0.25) | <0.01 | -0.46 | (-0.76, -0.17) | <0.01 |
| Within 5 km | < 70 | -0.05 | (-0.25, 0.15) | 0.62 | -0.26 | (-0.45, -0.06) | 0.01 | -0.09 | (-0.29, 0.11) | 0.39 | -0.13 | (-0.29, 0.02) | 0.09 |
| | 70-74 | 0.29 | (-0.25, 0.82) | 0.29 | -0.17 | (-0.69, 0.36) | 0.53 | -0.001 | (-0.51, 0.51) | 0.90 | 0.04 | (-0.37, 0.45) | 0.85 |
| | 75-79 | -0.78 | (-1.26, -0.31) | <0.01 | -0.72 | (-1.33, -0.11) | 0.02 | -0.46 | (-0.95, 0.03) | 0.06 | -0.65 | (-1.06, -0.25) | <0.01 |
| | ≥ 80 | -0.03 | (-0.60, 0.53) | 0.91 | -0.19 | (-0.82, 0.43) | 0.55 | -0.09 | (-0.70, 0.53) | 0.78 | -0.10 | (-0.61, 0.40) | 0.69 |
| **Fully adjusted**[2] | | | | | | | | | | | | | |
| Continuous (per 5 km) | < 70 | 0.02 | (-0.11, 0.14) | 0.78 | 0.05 | (-0.07, 0.17) | 0.37 | 0.02 | (-0.11, 0.14) | 0.80 | -0.03 | (-0.12, 0.06) | 0.54 |
| | 70-74 | -0.10 | (-0.51, 0.31) | 0.63 | 0.003 | (-0.39, 0.39) | 0.99 | 0.08 | (-0.32, 0.47) | 0.70 | 0.007 | (-0.30, 0.31) | 0.97 |

| | | | | | | | | | | | | | |
|---|---|---|---|---|---|---|---|---|---|---|---|---|---|
| | 75-79 | 0.50 | (-0.12, 1.20) | 0.11 | 0.90 | (-0.76, 0.93) | 0.84 | -0.10 | (-0.73, 0.53) | 0.76 | -0.16 | (-0.68, 0.36) | 0.54 |
| | ≥ 80 | 0.26 | (-0.41, 0.92) | 0.45 | 0.69 | (-0.01, 1.40) | 0.05 | 0.53 | (-0.15, 1.20) | 0.13 | -0.49 | (-1.10, 0.08) | 0.09 |
| Within 1.5 km | < 70 | -0.04 | (-0.31, 0.22) | 0.75 | -0.04 | (-0.30, 0.21) | 0.74 | 0.10 | (-0.16, 0.36) | 0.45 | 0.004 | (-0.20, 0.20) | 0.97 |
| | 70-74 | -0.34 | (-0.79, 0.10) | 0.13 | 0.41 | (-0.01, 0.83) | 0.06 | -0.39 | (-0.81, 0.03) | 0.07 | -0.11 | (-0.43, 0.22) | 0.51 |
| | 75-79 | -0.005 | (-0.57, 0.56) | 0.99 | -0.38 | (-1.14, 0.37) | 0.32 | 0.08 | (-0.49, 0.65) | 0.79 | -0.10 | (-0.57, 0.37) | 0.67 |
| | ≥ 80 | -0.24 | (-0.75, 0.26) | 0.35 | -0.78 | (-1.31, -0.25) | <0.01 | -0.33 | (-0.85, 0.19) | 0.22 | -0.45 | (-0.88, -0.02) | 0.04 |
| Within 3 km | < 70 | -0.07 | (-0.24, 0.10) | 0.42 | -0.17 | (-0.34, -0.01) | 0.04 | -0.02 | (-0.18, 0.15) | 0.84 | -0.09 | (-0.22, 0.04) | 0.18 |
| | 70-74 | -0.03 | (-0.38, 0.32) | 0.86 | 0.24 | (-0.09, 0.57) | 0.15 | -0.22 | (-0.55, 0.11) | 0.19 | -0.004 | (-0.26, 0.25) | 0.98 |
| | 75-79 | -0.01 | (-0.39, 0.37) | 0.97 | 0.33 | (-0.18, 0.84) | 0.20 | -0.02 | (-0.40, 0.36) | 0.93 | 0.10 | (-0.21, 0.42) | 0.53 |
| | ≥ 80 | -0.26 | (-0.64, 0.11) | 0.17 | -0.56 | (-0.95, -0.16) | 0.01 | -0.61 | (-0.98, -0.23) | <0.01 | -0.48 | (-0.79, -0.16) | <0.01 |
| Within 5 km | < 70 | 0.03 | (-0.19, 0.25) | 0.81 | -0.22 | (-0.43, -0.01) | 0.04 | 0.06 | (-0.16, 0.27) | 0.61 | -0.05 | (-0.21, 0.12) | 0.58 |
| | 70-74 | 0.30 | (-0.24, 0.84) | 0.28 | -0.21 | (-0.73, 0.31) | 0.42 | 0.05 | (-0.46, 0.57) | 0.84 | 0.05 | (-0.35, 0.44) | 0.82 |
| | 75-79 | -0.54 | (-1.05, -0.03) | 0.04 | -0.69 | (-1.37, -0.005) | 0.05 | -0.16 | (-0.68, 0.36) | 0.54 | -0.46 | (-0.88, -0.04) | 0.03 |
| | ≥ 80 | 0.05 | (-0.60, 0.70) | 0.88 | 0.19 | (-0.51, 0.89) | 0.60 | 0.44 | (-0.23, 1.10) | 0.20 | 0.22 | (-0.34, 0.79) | 0.43 |

[1]Minimally adjusted for baseline age and sex. [2]Fully adjusted for age, sex, education, income, marital status, race/ethnicity, smoking status, alcohol consumption. Continuous distance interpreted per every 5 km closer a residence is to a lead facility. β: beta value for linear regression estimate, km: kilometers, STAR: Study of Healthy Aging in African Americans.

**Supplemental Table 12:** Sensitivity results for education stratified analysis adjusted linear regression associations between residential distance to lead releasing facility and baseline cognition in KHANDLE analytic sample (n=1,638). Cognitive measures are z-score standardized.

| Distance to lead facility | Education level | Cognitive measure | | | | | | | | | | | |
|---|---|---|---|---|---|---|---|---|---|---|---|---|---|
| | | Episodic memory | | | Semantic memory | | | Executive function | | | Global cognition | | |
| | | β | 95% CI | P value | β | 95% CI | P value | β | 95% CI | P value | β | 95% CI | P value |
| **Minimally adjusted[1]** | | | | | | | | | | | | | |
| Continuous (per 5 km) | ≤ high school | -0.10 | (-0.03, -0.17) | <0.01 | -0.17 | (-0.10, -0.24) | <0.01 | -0.07 | (-0.0002, -0.13) | 0.05 | -0.11 | (-0.06, -0.17) | <0.01 |
| | trade school or college | -0.07 | (-0.03, -0.11) | <0.01 | -0.08 | (-0.04, -0.12) | <0.01 | -0.05 | (-0.01, -0.09) | 0.01 | -0.07 | (-0.04, -0.10) | <0.01 |
| | graduate school | -0.06 | (-0.13, 0.01) | 0.10 | -0.02 | (-0.09, 0.05) | 0.67 | 0.05 | (-0.02, 0.12) | 0.19 | -0.01 | (-0.06, 0.05) | 0.76 |
| Within 1.5 km | ≤ high school | -0.11 | (-0.52, 0.30) | 0.58 | -0.32 | (-0.75, 0.12) | 0.15 | -0.20 | (-0.59, 0.20) | 0.33 | -0.21 | (-0.54, 0.12) | 0.22 |
| | trade school or college | -0.17 | (-0.46, 0.11) | 0.24 | -0.17 | (-0.46, 0.13) | 0.26 | -0.35 | (-0.63, -0.06) | 0.02 | -0.23 | (-0.46, -0.002) | 0.05 |
| | graduate school | 0.54 | (-0.30, 1.39) | 0.21 | -0.07 | (-0.94, 0.80) | 0.87 | -0.10 | (-0.99, 0.80) | 0.83 | 0.13 | (-0.55, 0.80) | 0.71 |
| Within 3 km | ≤ high school | -0.29 | (-0.52, -0.05) | 0.02 | -0.38 | (-0.62, -0.13) | <0.01 | -0.37 | (-0.60, -0.15) | <0.01 | -0.35 | (-0.53, -0.16) | <0.01 |
| | trade school or college | -0.14 | (-0.28, -0.01) | 0.04 | -0.17 | (-0.30, -0.03) | 0.02 | -0.15 | (-0.28, -0.02) | 0.03 | -0.15 | (-0.26, -0.05) | <0.01 |
| | graduate school | -0.15 | (-0.41, 0.11) | 0.26 | -0.36 | (-0.63, -0.10) | 0.01 | -0.06 | (-0.34, 0.21) | 0.65 | -0.19 | (-0.40, 0.01) | 0.07 |
| Within 5 km | ≤ high school | -0.37 | (-0.57, -0.17) | <0.01 | -0.55 | (-0.76, -0.34) | <0.01 | -0.34 | (-0.54, -0.15) | <0.01 | -0.42 | (-0.58, -0.26) | <0.01 |
| | trade school or college | -0.24 | (-0.35, -0.13) | <0.01 | -0.24 | (-0.35, -0.13) | <0.01 | -0.15 | (-0.26, -0.04) | 0.01 | -0.21 | (-0.30, -0.12) | <0.01 |
| | graduate school | -0.11 | (-0.29, 0.06) | 0.21 | -0.04 | (-0.22, 0.14) | 0.66 | 0.05 | (-0.13, 0.24) | 0.59 | -0.03 | (-0.17, 0.10) | 0.63 |
| **Fully adjusted[2]** | | | | | | | | | | | | | |

| | | | | | | | | | | | | | |
|---|---|---|---|---|---|---|---|---|---|---|---|---|---|
| Continuous (per 5 km) | ≤ high school | -0.08 | (-0.01, -0.15) | 0.02 | -0.10 | (-0.04, -0.17) | <0.01 | -0.01 | (-0.08, 0.05) | 0.75 | -0.07 | (-0.01, -0.12) | 0.01 |
| | trade school or college | -0.04 | (-0.004, -0.08) | 0.03 | -0.001 | (-0.04, 0.03) | 0.95 | 0.002 | (-0.03, 0.04) | 0.90 | -0.01 | (-0.04, 0.01) | 0.33 |
| | graduate school | -0.04 | (-0.11, 0.02) | 0.20 | -0.01 | (-0.07, 0.05) | 0.65 | 0.04 | (-0.03, 0.10) | 0.24 | -0.01 | (-0.05, 0.04) | 0.79 |
| Within 1.5 km | ≤ high school | -0.01 | (-0.41, 0.39) | 0.95 | -0.33 | (-0.70, 0.03) | 0.08 | -0.14 | (-0.52, 0.23) | 0.45 | -0.16 | (-0.46, 0.14) | 0.29 |
| | trade school or college | -0.13 | (-0.41, 0.15) | 0.37 | -0.05 | (-0.30, 0.21) | 0.72 | -0.23 | (-0.48, 0.03) | 0.08 | -0.13 | (-0.34, 0.07) | 0.19 |
| | graduate school | 0.36 | (-0.46, 1.17) | 0.39 | -0.20 | (-0.91, 0.50) | 0.57 | -0.19 | (-0.97, 0.59) | 0.63 | -0.01 | (-0.57, 0.55) | 0.97 |
| Within 3 km | ≤ high school | -0.19 | (-0.44, 0.05) | 0.13 | -0.19 | (-0.41, 0.03) | 0.1 | -0.20 | (-0.43, 0.02) | 0.08 | -0.19 | (-0.38, -0.01) | 0.04 |
| | trade school or college | -0.06 | (-0.20, 0.07) | 0.37 | -0.0007 | (-0.12, 0.12) | 0.99 | -0.01 | (-0.14, 0.11) | 0.82 | -0.03 | (-0.12, 0.07) | 0.60 |
| | graduate school | -0.09 | (-0.35, 0.17) | 0.48 | -0.27 | (-0.50, -0.05) | 0.02 | 0.03 | (-0.22, 0.27) | 0.84 | -0.11 | (-0.29, 0.06) | 0.21 |
| Within 5 km | ≤ high school | -0.29 | (-0.51, -0.07) | 0.01 | -0.36 | (-0.56, -0.15) | <0.01 | -0.14 | (-0.35, 0.07) | 0.18 | -0.26 | (-0.43, -0.10) | <0.01 |
| | trade school or college | -0.16 | (-0.27, -0.05) | <0.01 | -0.03 | (-0.13, 0.07) | 0.56 | 0.002 | (-0.10, 0.11) | 0.97 | -0.06 | (-0.14, 0.02) | 0.12 |
| | graduate school | -0.06 | (-0.24, 0.12) | 0.51 | -0.01 | (-0.16, 0.15) | 0.93 | 0.06 | (-0.11, 0.22) | 0.52 | -0.004 | (-0.12, 0.12) | 0.95 |

[1]Minimally adjusted for baseline age and sex. [2]Fully adjusted for age, sex, education, income, marital status, race/ethnicity, smoking status, alcohol consumption. Continuous distance interpreted per every 5 km closer a residence is to a lead facility. β: beta value for linear regression estimate, km: kilometers, KHANDLE: Kaiser Healthy Aging and Diverse Life Experiences Study.

**Supplemental Table 13:** Sensitivity results for education stratified analysis adjusted linear regression associations between residential distance to lead releasing facility and baseline cognition in STAR analytic sample (n=741). Cognitive measures are z-score standardized.

| Distance to lead facility | Education level | Cognitive measure | | | | | | | | | | | |
|---|---|---|---|---|---|---|---|---|---|---|---|---|---|
| | | Episodic memory | | | Semantic memory | | | Executive function | | | Global cognition | | |
| | | β | 95% CI | P value | β | 95% CI | P value | β | 95% CI | P value | β | 95% CI | P value |
| **Minimally adjusted** [1] | | | | | | | | | | | | | |
| Continuous (per 5 km) | ≤ high school | -0.14 | (-0.61, 0.32) | 0.55 | -0.18 | (-0.70, 0.34) | 0.50 | -0.23 | (-0.69, 0.23) | 0.33 | -0.18 | (-0.55, 0.18) | 0.33 |
| | trade school or college | -0.09 | (-0.27, 0.09) | 0.35 | -0.15 | (-0.33, 0.04) | 0.12 | -0.05 | (-0.22, 0.12) | 0.56 | -0.09 | (-0.23, 0.04) | 0.18 |
| | graduate school | 0.002 | (-0.14, 0.15) | 0.97 | 0.03 | (-0.12, 0.17) | 0.73 | 0.02 | (-0.11, 0.16) | 0.76 | 0.02 | (-0.09, 0.12) | 0.77 |
| Within 1.5 km | ≤ high school | -0.18 | (-0.53, 0.17) | 0.32 | -0.21 | (-0.59, 0.18) | 0.30 | -0.19 | (-0.53, 0.16) | 0.28 | -0.19 | (-0.47, 0.08) | 0.17 |
| | trade school or college | -0.08 | (-0.32, 0.16) | 0.53 | 0.06 | (-0.19, 0.30) | 0.64 | 0.04 | (-0.19, 0.27) | 0.74 | 0.01 | (-0.18, 0.19) | 0.95 |
| | graduate school | -0.27 | (-1.10, 0.56) | 0.53 | -0.35 | (-1.17, 0.48) | 0.41 | 0.25 | (-0.51, 1.01) | 0.52 | -0.12 | (-0.73, 0.49) | 0.70 |
| Within 3 km | ≤ high school | -0.07 | (-0.35, 0.21) | 0.62 | -0.20 | (-0.5, 0.11) | 0.21 | -0.28 | (-0.55, -0.01) | 0.04 | -0.18 | (-0.40, 0.04) | 0.10 |
| | trade school or college | -0.07 | (-0.22, 0.09) | 0.40 | -0.06 | (-0.22, 0.10) | 0.45 | -0.08 | (-0.22, 0.07) | 0.31 | -0.07 | (-0.18, 0.05) | 0.26 |
| | graduate school | -0.07 | (-0.37, 0.24) | 0.68 | -0.005 | (-0.31, 0.30) | 0.98 | 0.01 | (-0.27, 0.29) | 0.94 | -0.02 | (-0.25, 0.21) | 0.86 |
| Within 5 km | ≤ high school | -0.04 | (-0.53, 0.45) | 0.87 | -0.10 | (-0.64, 0.44) | 0.71 | -0.19 | (-0.67, 0.28) | 0.42 | -0.11 | (-0.50, 0.27) | 0.57 |
| | trade school or college | -0.0050 | (-0.21, 0.20) | 0.97 | -0.24 | (-0.45, -0.03) | 0.03 | 0.01 | (-0.19, 0.21) | 0.95 | -0.08 | (-0.24, 0.08) | 0.34 |
| | graduate school | -0.11 | (-0.46, 0.23) | 0.52 | -0.10 | (-0.44, 0.24) | 0.57 | 0.05 | (-0.27, 0.36) | 0.76 | -0.05 | (-0.31, 0.20) | 0.67 |
| **Fully adjusted** [2] | | | | | | | | | | | | | |

| | | | | | | | | | | | | | |
|---|---|---|---|---|---|---|---|---|---|---|---|---|---|
| Continuous (per 5 km) | ≤ high school | -0.03 | (0.48, -0.54) | 0.90 | -0.33 | (-0.87, 0.21) | 0.23 | -0.28 | (-0.76, 0.20) | 0.25 | -0.21 | (0.17, -0.60) | 0.28 |
| | trade school or college | -0.04 | (0.16, -0.23) | 0.72 | -0.14 | (-0.34, 0.06) | 0.16 | 0.04 | (-0.15, 0.22) | 0.70 | -0.05 | (0.10, -0.20) | 0.53 |
| | graduate school | 0.002 | (-0.15, 0.15) | 0.98 | 0.02 | (-0.12, 0.17) | 0.74 | 0.02 | (-0.12, 0.15) | 0.82 | 0.01 | (0.12, -0.09) | 0.80 |
| Within 1.5 km | ≤ high school | -0.84 | (-2.60, 0.92) | 0.35 | -1.69 | (-3.55, 0.17) | 0.07 | -1.07 | (-2.73, 0.59) | 0.21 | -1.20 | (-2.52, 0.13) | 0.08 |
| | trade school or college | -0.06 | (-0.31, 0.18) | 0.62 | 0.08 | (-0.17, 0.33) | 0.52 | 0.07 | (-0.16, 0.30) | 0.54 | 0.03 | (-0.16, 0.21) | 0.75 |
| | graduate school | -0.26 | (-1.10, 0.58) | 0.55 | -0.38 | (-1.21, 0.45) | 0.37 | 0.24 | (-0.54, 1.01) | 0.55 | -0.13 | (-0.75, 0.48) | 0.67 |
| Within 3 km | ≤ high school | 0.02 | (-0.28, 0.31) | 0.92 | -0.24 | (-0.55, 0.07) | 0.13 | -0.29 | (-0.57, -0.02) | 0.03 | -0.17 | (-0.39, 0.05) | 0.13 |
| | trade school or college | -0.04 | (-0.21, 0.12) | 0.60 | -0.06 | (-0.23, 0.11) | 0.48 | -0.03 | (-0.19, 0.13) | 0.70 | -0.05 | (-0.17, 0.08) | 0.48 |
| | graduate school | -0.14 | (-0.48, 0.20) | 0.42 | 0.02 | (-0.32, 0.35) | 0.92 | 0.04 | (-0.27, 0.36) | 0.78 | -0.03 | (-0.27, 0.22) | 0.84 |
| Within 5 km | ≤ high school | 0.12 | (-0.46, 0.69) | 0.69 | -0.15 | (-0.76, 0.46) | 0.63 | -0.2 | (-0.74, 0.35) | 0.48 | -0.08 | (-0.51, 0.36) | 0.73 |
| | trade school or college | 0.06 | (-0.17, 0.28) | 0.61 | -0.25 | (-0.48, -0.03) | 0.03 | 0.11 | (-0.10, 0.32) | 0.32 | -0.03 | (-0.20, 0.14) | 0.74 |
| | graduate school | -0.18 | (-0.56, 0.21) | 0.37 | -0.06 | (-0.44, 0.32) | 0.76 | 0.11 | (-0.24, 0.46) | 0.55 | -0.04 | (-0.32, 0.24) | 0.76 |

[1]Minimally adjusted for baseline age and sex. [2]Fully adjusted for age, sex, education, income, marital status, race/ethnicity, smoking status, alcohol consumption. Continuous distance interpreted per every 5 km closer a residence is to a lead facility. β: beta value for linear regression estimate, km: kilometers, STAR: Study of Healthy Aging in African Americans.

**Supplemental Figure 3.** Forest plot of mean difference 95% confidence intervals for minimally adjusted models in the KHANDLE and STAR analytic samples, and meta-analyzed across cohorts.

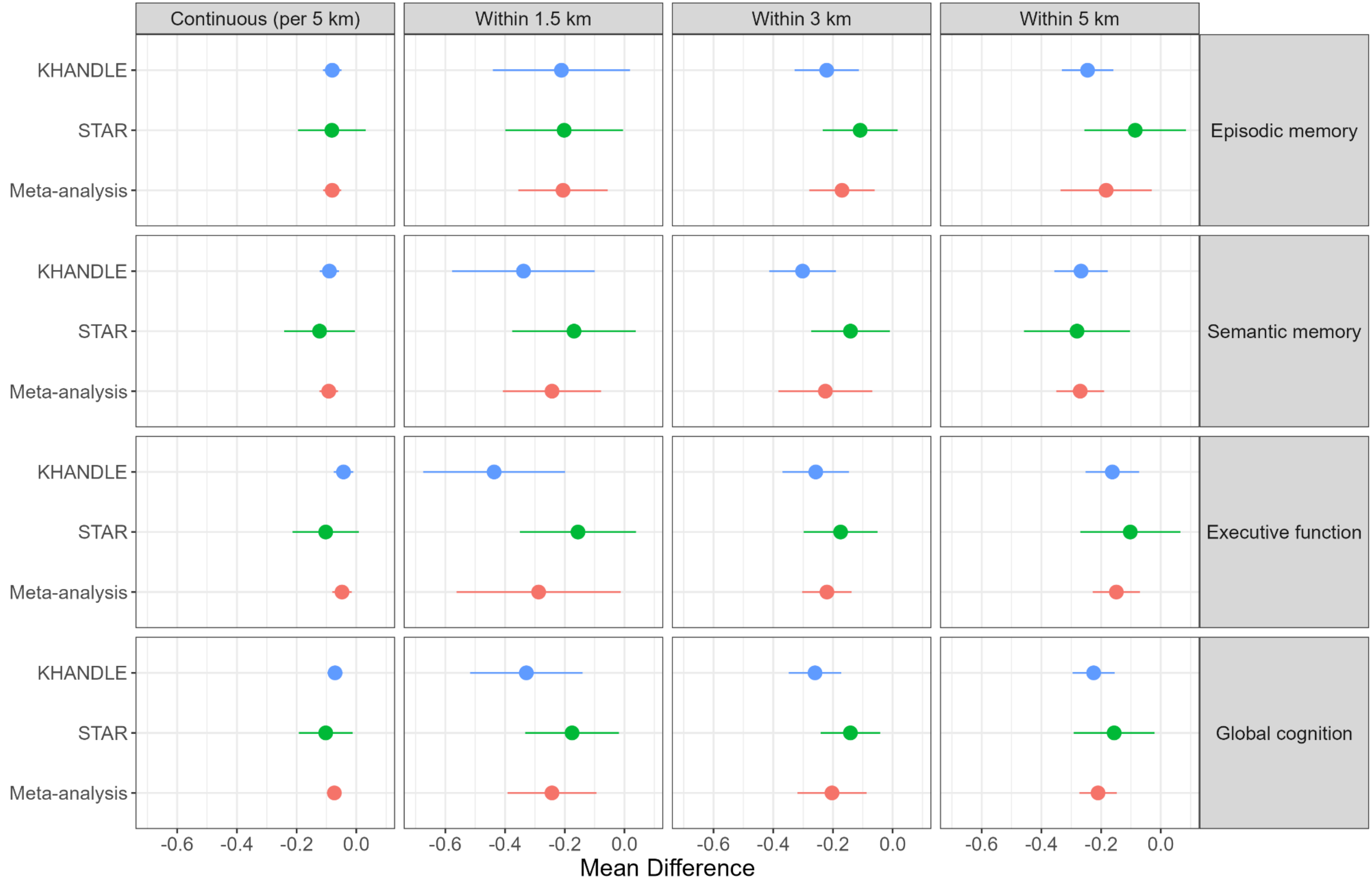


Minimally adjusted for age and sex. KHANDLE: Kaiser Healthy Aging and Diverse Life Experiences Study; STAR: Study of Healthy Aging in African Americans
